# The burden of rheumatic heart disease and issues affecting the provision of care in Malawi: A scoping review

**DOI:** 10.1101/2025.02.04.25321685

**Authors:** E Blennerhassett, O Brady Bates, M O’Connor, H Gondwe, L Msimuko, M Ledwidge, B Mbakaya, J Gallagher

## Abstract

**Background:** Rheumatic heart disease (RHD) is an autoimmune sequela of group A streptococcal (GAS) pharyngitis. Single or recurrent episodes of acute rheumatic fever (ARF), a complication arising 2-3 weeks after GAS infection, can cause damage to the valves of the heart and RHD. This trajectory towards severe disease is now rare in high-income countries. The appropriate use of antibiotics has contributed to this decline in RHD prevalence, but socio-economic factors have also had a profound impact. In Malawi, as in many low-income countries, RHD continues to pose a significant challenge. Limited access to healthcare and poor education likely contribute to the disease burden, and actionable solutions are needed to eradicate RHD in these communities.

**Aim:** To determine the present burden of GAS, ARF and RHD in Malawi, the issues affecting the provision of care and the solutions that have been proposed.

**Design and setting:** A scoping review encompassing four electronic databases from which relevant articles were drawn.

**Method:** A systematic search of ‘PubMed’, ‘EMBASE’, ‘Cochrane Library’ and ‘Clinicaltrials.gov’ was conducted to identify research related to the study objectives. Supplemental grey literature and internet searches were carried out and collaboration with experts in the field ensured a comprehensive review of the available data. The selected articles spanned from 2008 to October 2024 and addressed one or more of the diseases of interest. A scoping review was carried out according to the Arksey and O’Malley procedure.

**Results:** This scoping review includes thirty papers, which focus primarily on RHD burden, screening, treatment, and barriers to care in Malawi. There was significantly less research on ARF and GAS infections. A wide variety of study designs were employed (mostly descriptive, cross-sectional and retrospective studies) and the number of participants varied significantly across the different studies. Two government reports were also included. The primary findings included a shift in the focus of research towards solutions to the high burden of RHD in Malawi.

**Conclusion:** The significant morbidity and mortality associated with RHD are a major concern in the communities and healthcare systems of Malawi. As in many low-income countries, resource mobilisation and improved education are areas requiring attention. To address the high burden of disease in the country, ongoing research is largely focused on establishing a sufficiently large and appropriately trained workforce to diagnose and monitor RHD using the resources available within the constraints of the country’s socioeconomic context.

## Introduction

Rheumatic heart disease (RHD) is an acquired heart disease resulting from repeated infections with Group A Streptococcus (GAS), a bacteria that causes pharyngitis and cellulitis. GAS infection can trigger an autoimmune response leading to rheumatic fever (RF) which targets the valves of the heart, damaging them. Recurrent episodes of RF can evolve into RHD, which is characterised by permanent valvular damage. This progression has serious implications for cardiac function, leading to heart failure and arrhythmias, and significant morbidity and mortality if left untreated (1). The prevalence of RHD is markedly lower in high-income countries, inversely related to a country’s income status (2). Malawi, one of the world’s poorest nations, had an estimated 323,422 RHD cases in 2021 (3, 4). The persistence of RHD in the region is attributed to a complex interplay of factors including overcrowded living conditions, inadequate sanitation, poor nutrition and limited access to healthcare and education(5–7).

In contrast to high-income countries where GAS can be readily detected using throat swabs and rapid antigen tests, diagnoses in resource-constrained settings rely primarily on clinical judgement(8). The diagnostic role of echocardiography emerges as repeated episodes of ARF incur significant damage to the heart’s valves, causing RHD. There has been widespread adoption of portable echocardiography across Malawi since the 1990s, and its value in RHD care has been well-established in the literature from the country in the interim(9). RHD screening programmes employ echocardiography to examine cardiac function and anatomy at the point of care and ideally detect “latent” RHD. This is an asymptomatic stage of disease in individuals with no known ARF history but with pathologic changes to the heart confirmed on echocardiography. The echocardiography findings are graded according to the World Heart Federation guidelines as ‘normal’, ‘borderline RHD’ or ‘definite RHD’ with further subdivisions according to the valvular pattern of disease(10). In high-income countries, such screening programmes have facilitated timelier interventions and the virtual eradication of RHD. Limited access to the necessary equipment and individuals trained in their use have impeded similar progress in low-income countries.

RHD is a severe complication of treatable preceding infections. Primary prevention involves the institution of antimicrobial therapy for patients presenting with a confirmed or suspected GAS infection(1). Prompt diagnosis is particularly important for those at risk of developing further GAS infections or ARF(11), with the greatest risk attributed to overcrowding in the household environment, increasing the risk for lower-income communities(8). Current evidence suggests that the most effective secondary prevention strategy is the prevention of rheumatic fever with monthly intramuscular injections of benzathine penicillin G (BPG) over the course of many years(1). The associated requirement for consistent patient monitoring poses a substantial challenge, particularly for those living in poverty, further highlighting the need to promote RHD screening and early treatment (12).

However, despite these proven preventative measures, research from Malawi suggests that the majority of RHD cases present late, with severe symptomatic disease. At this stage, patients are less likely to achieve adequate disease control with BPG injections alone (10, 13). In these communities, the challenges to treatment become increasingly difficult to surmount as the disease progresses. The definitive treatment for advanced RHD is cardiac surgery, access to which is virtually absent in most low-to middle-income countries. At present, there are no options for surgical intervention for RHD in Malawi and cost-effective solutions and alternatives will be critical to addressing the high prevalence in the country(5).

The purpose of this scoping review is to comprehensively document existing RHD research in Malawi and to identify knowledge gaps. Through this approach, the study seeks to guide future research focused on bridging the gap between high- and low-income countries in the pursuit of RHD eradication.

## Objectives

The aim of this scoping review is to explore the available literature and to present a summarised log of the same.

### Methods

A scoping review was deemed the most appropriate approach to determine the burden of GAS, ARF and RHD in Malawi, and the obstacles to adequate treatment for these illnesses. Scoping reviews broadly survey the existing literature, allowing the prevailing themes and gaps in the research to be identified. This scoping review was designed as per the six-stage protocol outlined by Arksey and O’Malley (2005) and was conducted as follows (14):

#### 1. Identifying the research question

Although readily treatable through primary and secondary prevention strategies, RHD continues to cause significant morbidity and mortality in much of the developing world. The objective of this scoping review is to understand the following research question: *what does the available literature tell us about the burden of RHD, ARF and GAS infection in Malawi and what barriers impede care for affected populations?*

To define the burden of GAS, ARF and RHD in Malawi, the following outcomes of interest will be investigated:

- Incidence of sore throat in children aged 5-15 years
- Prevalence of GAS among cases of sore throat
- Incidence of ARF
- Case-fatality rate from ARF
- Prevalence of RHD
- Mortality from RHD
- Prevalence of non-fatal outcomes of RHD

To establish what barriers and facilitators exist in the detection and treatment of GAS, ARF and RHD, the following will be considered:

- Initial decision to seek care
- Factors influencing diagnosis
- Factors influencing treatment or referral
- Factors influencing adherence and retention in long-term care

The role of the patient, healthcare provider and health system in each of these will be examined.

#### 2. Identifying relevant studies

The databases included in the initial search were: PubMed, EMBASE, Cochrane Library, and Clinicaltrials.gov. Keywords were identified and used to establish medical subheadings (MeSH) and Emtree terms in Medline and EMBASE, respectively. A number of search terms were utilised to create a reading list. The search terms were grouped and eligible studies included one term from each group. A search strategy was generated and applied using a combination of terms linked to rheumatic heart disease, rheumatic fever and pharyngitis. The search strategies are available in the Supplemental Information (S2). Date (1995-2024), language (English) and country (Malawi) filters were applied. Further sources were identified by manually searching the reference lists of studies selected from the databases of interest.

#### 3. Study selection

The studies chosen for inclusion in the current review satisfied the following criteria: any epidemiological study (i) focused on the individual presenting with sore throat/ARF/RHD and identifying the principal stakeholders; (ii) addressing at least one of the diseases of interest (GAS, ARF, RHD); (iii) containing one or more of the objectives outlined in 1. the research question above; (iv) conducted in or after 1995. The exclusion of research published prior to 1995 ensured that the data reflected advances in the diagnosis and care of RHD (echocardiography was only widely implemented for this purpose post-1990s)(9). Publications including participants of all ages and genders in all healthcare settings were considered. The scope of the search was restricted to literature written in English. Exclusion criteria included opinion pieces, case reports and systematic reviews.

**Table 1.** Study inclusion and exclusion criteria.

| Inclusion criteria | Exclusion criteria |
| --- | --- |
| Published 1995-2024 | Published prior to 1995 |
| Published in English | Published in languages other than English |
| Focused on the individual presenting with GAS, ARF or RHD | Not focused on the individual presenting with GAS, ARF or RHD |
| Journal articles, government documents, theses from Malawi | Opinion pieces, case reports, systematic reviews |

Below is an outline of the Preferred Reporting Items for Systematic Reviews and Meta-Analyses (PRISMA) flow chart to determine the eligibility of individual studies found in the search for this scoping review.

#### 4. Charting the data

From the bank of relevant articles (n= 30), the following information was charted;

- First author
- Year of publication
- Literature type
- Type of study
- Area evaluated
- Main findings

#### 5. Collating, summarising and reporting results

A summary of the literature and relevant findings can be found in the Supplemental Information (S1). These findings are further elaborated in the results section.

#### 6. Consultation exercise

The inclusion and exclusion of studies from this scoping review were guided by the advice of experts in the field of RHD in Malawi.

## Results

### Description of included studies

The PRISMA ScR flowchart (Fig 1) above outlines the approach employed for searching, identifying and selecting the relevant records. The preliminary searches of PubMed, EMBASE, Cochrane Library and Clinicaltrials.gov yielded thirty-five records. An additional nine papers were identified by hand-searching the reference lists. Following a review of the study titles and abstracts and the removal of duplicates, thirty records met the eligibility criteria and were deemed appropriate for full-text review. At the end of this process, all thirty papers were accepted for data extraction and inclusion in the scoping review. Ten records included were conference abstracts, and seventeen were articles published in peer-reviewed journals. One clinical trial was included. The two remaining records were government policy documents included on the recommendation of an expert in the field of RHD in Malawi.

**Fig 1.**
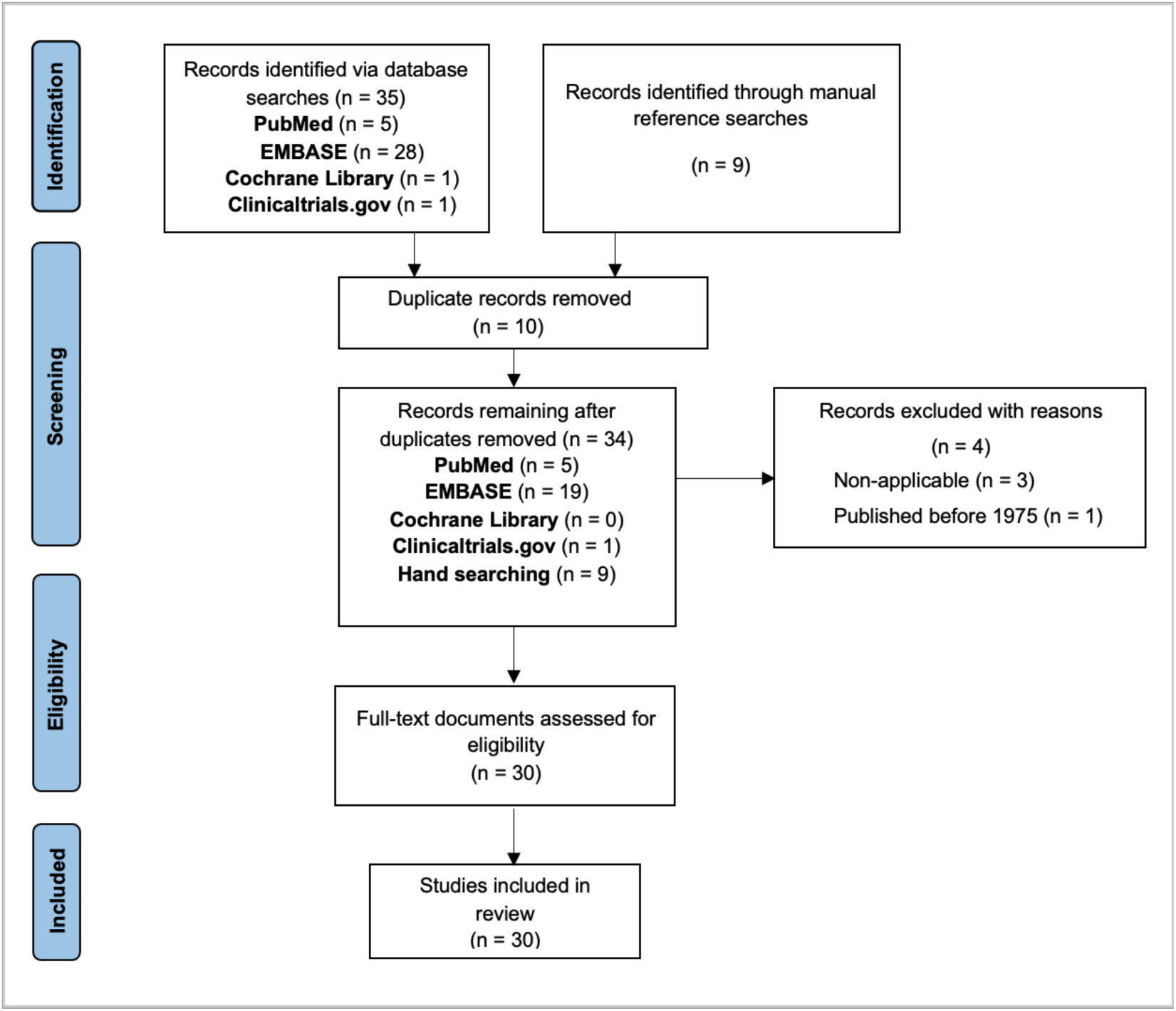
**PRISMA ScR flow-chart.**

### Study design

The papers included in this review employed a wide variety of study designs. Among the thirty records included, there were six descriptive studies (6, 15–19), four cross-sectional studies (20–23), four cohort studies (12, 13, 24, 25), two pilot studies (5, 26), two retrospective analyses (27, 28), one observational study(29), one feasibility study (30), one concept design (31), one scoping review (32), one literature review (9), one secondary analysis (33), one external validation study (34), one interventional study (35), and one mixed-method study combining qualitative and quantitative strategies (36). One clinical trial and two government report documents were also deemed important for inclusion in the review (37–40).

### Study population and setting

The study sample sizes varied significantly, ranging from 1,450 to 3,908 participants in Malawi-specific RHD screening studies (12, 28). RHD participant’s ages varied from 5-16 (20, 34) in studies from Malawi and there was a female preponderance. Most studies were based in urban and peri-urban settings. Seven studies were carried out in central hospitals; four in Kamuzu Central Hospital in Lilongwe (5, 6, 24, 29), two in Queen Elizabeth Central Hospital in Blantyre (18, 41), and one in Mzuzu Central Hospital (28). Three were conducted in schools and/or nearby communities in Lilongwe (17, 20, 21) and Blantyre (21). Two studies were based in rural PEN-Plus clinics, in Neno District Hospital and Lisungwi Community Hospital, offering decentralised care for RHD 110km from the nearest tertiary centre (16, 35). One study was based on Likoma Island (30).

### Outcomes of interest

#### The burden of GAS, ARF and RHD in Malawi

The burden of GAS infection in Malawi was not formally assessed in any of the papers included in this review, although two studies reported ARF prevalence data. Zühlke et al. found that within their cohort, 22.3% of RHD patients from low-income countries had a documented episode of ARF in their medical history(19). The data collected in this study were grouped according to the country’s economic status, and Malawi-specific figures could not be extrapolated. Another study reported that 13% of children diagnosed with RHD (n = 39) in their cohort had a history of ARF(6). Two large multi-centre studies found that the incidence of ARF recurrence was low during the study period, irrespective of the country’s income status(29). For instance, Zühlke et al. (2016) recorded a 0.6% incidence of ARF recurrence over a 27-month period assessing 14 low- and middle-income countries(25).

Several studies addressed the burden of RHD in Malawi according to the criteria of interest within this review including the incidence and prevalence of RHD, and the associated morbidity and mortality.

One study evaluated the incidence of subclinical RHD evolution (between normal, borderline and definite echocardiographic categories) in Malawian schoolchildren and found that the progression or regression of subclinical disease varied substantially across participants by the end of the study period. Factors associated with changes in disease status included the pattern of valvular involvement and the use of antibiotic prophylaxis (13).

Overall, six studies assessed the prevalence of RHD in Malawi; one focused on latent RHD in urban and peri-urban communities(20) and another investigated RHD prevalence in rural communities. Latent RHD was found to have a prevalence of 3.4% in schoolchildren living within 60km of the country’s capital city, Lilongwe, and the highest prevalence was among 11-13-year-olds (20). The rural prevalence of RHD in Malawi could not be discerned from the latter study as the data from multiple low-income countries was pooled for analysis (23). Two further studies investigated RHD prevalence in cardiology clinics in Malawi, one in a paediatric population and the other in adult patients. 22% of the children (n = 250) and 30% of the adults (n = 3908) presenting to the respective cardiology clinics had manifest RHD (28). In another study based on Likoma Island in Lake Malawi, 5.28% of the children who underwent echocardiographic screening (n = 416) had RHD (30).

A number of studies investigated the outcomes of RHD, in terms of mortality and morbidity. Fatal outcomes were evaluated in three studies (15, 25, 29). One study reported that 0.6% of inpatient deaths in a large tertiary hospital in Malawi were attributable to RHD or rheumatic fever, while the leading causes of death were sepsis and lower respiratory tract infections(15). Two additional studies combined data from several low-income countries for analysis, but Malawi-specific data were not independently reported(25, 29). Across these two studies, patient factors including increasing age, lower income status, poorer education and more severe valvular disease were associated with increased RHD mortality(25, 29). Karthikeyan et. al (2024) reported that, across income groups, heart failure and sudden cardiac death accounted for the majority of RHD-associated deaths(29). Mortality was lower among females, participants with an education beyond primary school, those with a history of valvuloplasty or valve surgery, and those using secondary antibiotic prophylaxis. While a smaller proportion of patients from low-income countries underwent valve surgery, the use of secondary antibiotic prophylaxis was higher in this cohort compared to those in middle-income countries(29).

Non-fatal outcomes and complications of RHD were investigated in four studies. The adverse clinical outcomes described included congestive heart failure (CHF), stroke, atrial fibrillation, infective endocarditis, and worsening echocardiographic RHD status(19, 25, 29, 34).

One study of RHD across multiple countries found that more patients in low-income countries were diagnosed with heart failure, both at baseline and the two-year follow-up, compared to those in upper-middle-income countries. Conversely, stroke and infective endocarditis were more prevalent in upper-middle-income groups(25). Another, more recent study that included Malawi reported that the incidence of stroke, infective endocarditis, and recurrent rheumatic fever was low across income groups during the three-year follow-up period. Supporting earlier findings, this study also found that the highest risk of stroke was in the upper-middle income group(29).

Patterns of disease evolution were evaluated in two studies (12, 13). Four studies evaluated the characteristics of RHD, highlighting the extent of severe forms of the disease (6, 21, 28, 42). Another two documented patient characteristics as well as complications of the disease (19, 25). Of note, a significant number of patients with RHD in Malawi present to healthcare late with severe disease. Severe first presentations were reported between 62%-82.5% across three separate studies (6, 17, 21).

### Barriers to care

#### Factors influencing the initial decision to seek care

Within the studies included in this review, there was limited information about the factors influencing an individual’s initial decision to seek care. The available research consistently reiterates the observation that RHD patients in Malawi present to healthcare with advanced disease (6). This is attributed, in part, to inadequate education about the dangers associated with repeated ARF episodes and the logistical obstacles to healthcare access.

The PEN-Plus clinics established in rural Neno have been instrumental, particularly for those living remotely, by reducing the need for long-distance travel and improving access to healthcare for the poorest communities in Malawi. These clinics are run by mid-level providers trained to diagnose RHD, implement early treatment plans, and refer to specialists where appropriate. Additionally, community health workers visit patients who miss follow-up clinics to identify and address barriers that may exist for the individual and to re-enrol them in care(16).

#### Factors influencing diagnosis

Many studies found that inadequate RHD-specific education for Malawian communities and healthcare workers poses a significant challenge to RHD diagnosis and management in the country. Recent research from Malawi is largely focused on the assessment of RHD education in the country and the implementation of programmes aimed at improving awareness.

Eight studies in this review investigated RHD education strategies, with an emphasis on training non-physician providers to diagnose and treat RHD (5, 16, 22, 26, 27, 30, 32, 35). This approach, frequently referred to as “task-shifting”, emerges as a salient theme across the literature. There was a consensus across these studies that training nurses and clinical officers, collectively termed mid-level providers, was a feasible and cost-effective means for improving RHD diagnosis in Malawi.

The structure of the education programmes varied across the different studies with courses spanning from several days to months and combining didactic lectures, supervised ultrasound training including image interpretation alongside clinical mentoring (26, 27, 35). Trainees were at various qualification levels and included medical students, general physicians, clinical officers, nurses and physician assistants. The number of trainees ranged from two to sixty-five(5, 30).

The evaluation of training efficacy also differed among these studies. Three studies examined the sensitivity and specificity of RHD diagnosis after training, finding reasonable congruence between trainees’ and cardiologist diagnoses (22, 26). Two studies conducted pre- and post-training assessments, with retention assessments one year and six months later, respectively. At six months, test scores remained high at an average of 84.4%, compared to a pre-test average of 63.3% and an immediate post-test average of 88.3%. In another study, average test scores one year after training had fallen to 69.5% from 83.2% in the immediate post-training period, highlighting a need for ongoing training and re-assessment to ensure that adequate standards are maintained among these providers (27, 35). Another study employed a post-workshop questionnaire.

One study assessed the standard of knowledge among healthcare providers regarding the prevention of ARF and RHD in a central hospital. Participating nurses and clinicians reported concerns about the safety of BPG for RHD prevention. A subsequent RHD education workshop improved participants’ understanding of and comfort with prescribing the medication (5).

#### Factors influencing treatment and adherence

Many studies in this review reveal that inadequate resources, in the form of medication, medical equipment and human power, impede RHD care in Malawi in a number of ways (12, 19, 29, 33). For instance, one study identified resource limitations as a key barrier to treatment adherence. After the first year of the study, the healthcare centres treating the participants did not have a steady supply of BPG. Therefore, many children did not receive their injections despite presenting for their appointments. In light of this, BPG was provided directly to the participants for the following year which improved adherence but revealed a further barrier to treatment; an insufficient supply of the syringes required for BPG administration at the healthcare centres (12). A large multi-centre study similarly found that a significant proportion of RHD patients in low-income countries (up to 50%) received suboptimal secondary RHD prophylaxis(19).

Alongside medication and equipment shortages in Malawi, the workforce is insufficiently powered to facilitate long-term follow-up and surgical interventions for many RHD patients. Follow-up is critical to ensure appropriate RHD prophylaxis, alongside monitoring for and control of RHD complications. As mentioned, “task-shifting” is likely to play a central role in expanding the effective workforce to improve longitudinal RHD care. Unfortunately, the lack of cardiac surgeons in the country also poses an issue. The REMEDY study revealed that valvular repair and replacement were positively correlated with a country’s income level (19). In this study population, there were twice as many valve surgeries carried out in high-income-relative to low-income-countries, despite a higher prevalence of RHD cases requiring surgical intervention in the latter. This finding was reiterated in a more recent multi-centre study, which reported that 3.3% of RHD patients across low-income countries underwent valve surgery compared to 7.2% of those from upper-middle-income countries included in the study. Karthikeyan et al. suggest that referring patients with advanced RHD to lower-middle-income countries like India for valve surgery may be more cost-effective than developing in-country surgical infrastructure in low-income countries like Malawi(29).

#### Factors influencing retention in long-term care

The factors influencing retention in long-term care were not elucidated in the studies in this review. An interplay between the prolonged nature of RHD treatment and various socio-economic factors is likely. One research group showed that by utilising established patient tracking systems in Malawi, such as the HIV-defaulter system, they could maximise retention in care throughout the study period providing a possible solution to the issue of long-term follow-up of RHD patients in Malawi(12).

## Discussion

This scoping review sought to outline the current literature on the burden of rheumatic heart disease in Malawi, the barriers to RHD care and the solutions that have been proposed. In 2015, a list of strategies for ARF and RHD elimination in Africa was outlined by RHD experts in collaboration with the Social Cluster of the African Union Commission. In Malawi, the body of research assessing these strategies, which include decentralisation of technical expertise for RHD diagnosis and the creation of disease registers, has steadily grown. Nonetheless, there remain numerous challenges to overcome which will require a better understanding of the barriers to care for individuals and communities, a sufficient supply of penicillin for those affected, cardiac surgery centres for treatment of advanced RHD and collaboration with international corporations to facilitate resource mobilisation, monitoring and programme evaluations (43).

### Key findings

#### Screening and diagnosis

The optimisation of RHD echo-based screening and diagnosis in Malawi is a prominent theme in this review. Undoubtedly, screening programmes have important diagnostic implications and have provided insight into the burden of RHD in Malawi(3, 5). However, widespread screening in endemic regions may also present challenges, such as the over-treatment of latent RHD. A recent randomised controlled trial revealed a comparably high rate of latent RHD regression in both BPG-treated and placebo groups, suggesting that screening may lead to unnecessary treatment in some individuals who are not expected to yield a clear benefit(44). The use of additional tools, such as simplified echocardiographic scoring systems for latent RHD, may minimise this. By stratifying patients according to their risk of progression, the limited supply of medications available can be appropriately allocated to those most at risk of unfavourable outcomes(34).

#### Rural health disparities

A notable gap in current research lies in understanding the true prevalence of RHD in rural communities, which account for 84% of Malawi’s population. Most of the studies in this review are based in urban or peri-urban settings, and many in the country’s largest hospitals(19, 28). The research to date, therefore, unlikely accounts for a large proportion of the population for whom tertiary care centres are not readily accessible (28, 37). RHD research from other countries in Sub-Saharan Africa indicates an increased disease prevalence in rural communities, suggesting that a substantial number of RHD cases in Malawi are being overlooked, possibly complicating eradication efforts(45, 46).

#### Antecedent illnesses research

There is a limited body of research from Malawi on the illnesses preceding RHD, namely GAS and ARF. While the relevant terminology may not have been captured in the search strategy, this finding may also reflect health-seeking behaviours in Malawi. The scarcity of research may reflect low rates of healthcare presentation for these early infections and a failure to capture them in studies conducted within the formal healthcare system. Conducting research at the community level to evaluate individuals’ decision to seek or avoid healthcare would enhance current research and provide a deeper understanding of the barriers to care (6). Furthermore, assessing the feasibility of primordial prevention strategies, aimed at minimising risk factors to prevent RHD development, will be crucial. Despite widespread recognition of the link between poor social conditions and GAS/ARF development, little is known about the impact that changes at this level may have on disease prevalence and trajectory in the resource-poor communities most susceptible to RHD (19).

#### RHD-specific education for Malawian healthcare workers

There has been a notable shift in the focus of RHD research from Malawi in recent years. While earlier studies laid crucial foundations by delineating disease patterns and emphasising the significant burden of disease, recent efforts have shifted towards addressing the shortage of physicians trained in RHD care. Task-shifting programmes, which educate non-physician healthcare workers in the diagnosis and treatment of RHD, offer promise. This model is currently in place at the PEN-Plus clinics in Malawi and similar models have shown success elsewhere, including in rural Rwanda, by reducing the need for specialist cardiology input in resource-poor settings(16, 47).

#### RHD-specific education for Malawian communities

Notably, there remains a dearth of research into education initiatives to increase RHD awareness within the community. Although relevant sources may have been erroneously omitted from the search strategy, this finding may point to a gap in the literature. In the context of poor healthcare education in the community the link, for example, between GAS-pharyngitis and serious long-term complications like RHD may not be widely known. As a result, many may progress from subclinical to manifest disease before seeking out care.

#### Retention in care

The retention of RHD patients in care long-term in Malawi is a substantial challenge, with multifactorial aetiology. Medication and equipment shortages, coupled with lower education status and the necessity for long-distance travel to healthcare facilities hinder consistent engagement with healthcare services (19, 25, 48, 49). This is highlighted by a large multi-centre RHD study, which reported an 11.5% loss of participants to follow-up, predominantly among individuals with severe disease or lower education status. These findings underscore the barriers to the continuity of care in impoverished communities (25).

Possible alternative treatments that mitigate the need for fortnightly hospital visits have not been investigated in these studies. However, research from the PEN-Plus clinics suggests a possible solution. Training community workers to conduct home visits and provide social support for those absent from follow-up appointments has improved patient engagement in these centres and a roll-out of these strategies across the country could considerably improve retention in care, ultimately improving patient outcomes and reducing the burden of RHD in Malawi(16).

## Strengths & limitations

By utilising the Arksey and O’Malley framework, this scoping review mapped the available RHD literature from Malawi according to a rigorous and standardised approach for study selection and analysis. Nonetheless, the scoping review process inherently lacks an assessment of study quality, instead outlining all relevant resources regardless of quality, unlike a systematic review. Additionally, the literature search was limited to articles written in or translated into English and the results of this scoping review are unlikely to represent the full repertoire of research available on the topic of RHD in Malawi. Finally, a number of studies included Malawian participants as part of a low-income group without presenting each country’s data separately, thus allowing only generalised deductions to be made in these instances.

## Comparisons with the existing literature

One other scoping review detailing RHD research in Malawi was identified in our study selection process. The findings of this scoping review build upon those of Zhao et. al, who mapped the literature for task-sharing in Malawi. In the interim, further studies on task-sharing in Malawi have been published, underscoring the importance of maximising the country’s available human resources in the efforts for RHD eradication. This review complements the existing literature by mapping the current research landscape, thus highlighting prevailing themes and identifying gaps that can guide future research in a meaningful way.

## Implications of this review

This scoping review’s findings suggest that the burden of RHD and, by extension, preceding illnesses remains high in Malawi despite the availability of cost-effective means of screening and treatment. A multitude of contributory factors are at play. At this time, Malawi has a single paediatric cardiologist and no in-country cardiac surgeons despite a population of 22 million and growing and despite a significant burden of cardiac disease in the country (16). Further issues identified include a failure to recognise and manage GAS pharyngitis/ARF in primary care (50), a lack of education for communities and healthcare workers about primary and secondary prevention, a high prevalence of asymptomatic disease which hinders health-seeking behaviours, and barriers to care including longitudinal monitoring with repeated BPG injections.

## Conclusion

In Malawi, and throughout Sub-Saharan Africa, RHD remains a leading cause of morbidity and mortality despite successful eradication efforts in high-income countries (51). If the WHO is to achieve its goal of a 25% reduction in mortality from rheumatic fever and RHD in under 25s by the year 2025, future research from Malawi, where the burden is significant, must provide direction to that end (52). Resource scarcity and additional barriers to care must be considered in the solution. The questions to be addressed are; why does RHD persist in these communities and what solutions are realistic in the socio-economic setting of countries like Malawi? The answers will guide population-specific RHD care, inform government policy and facilitate appropriate resource allocation, with the goal of reducing the significant health inequity between high- and low-income countries, such as Malawi.

## Data Availability

All data produced in the present work are contained in the manuscript

## Supplemental information

**S1 Table. Description of studies included.**

**Table S1.**
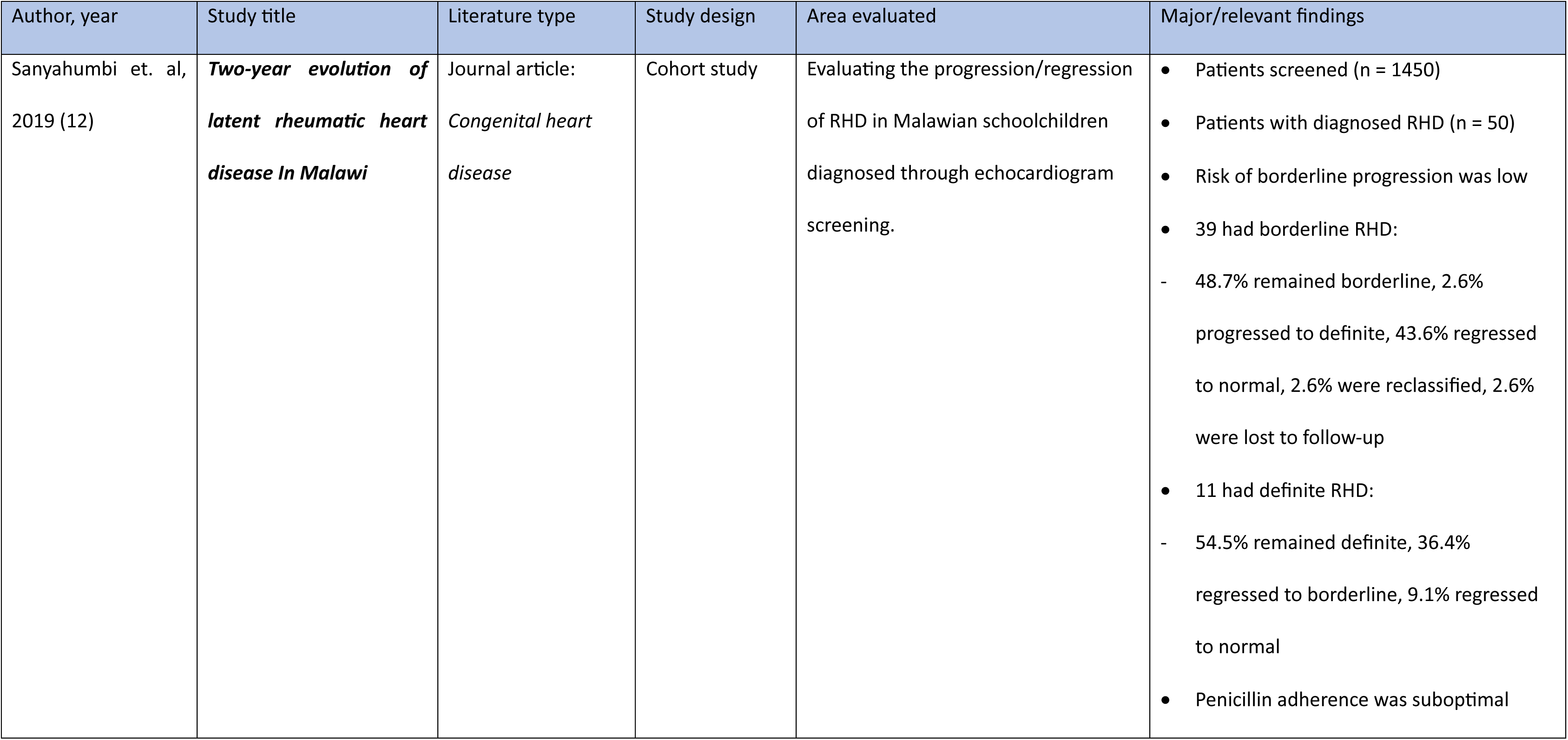
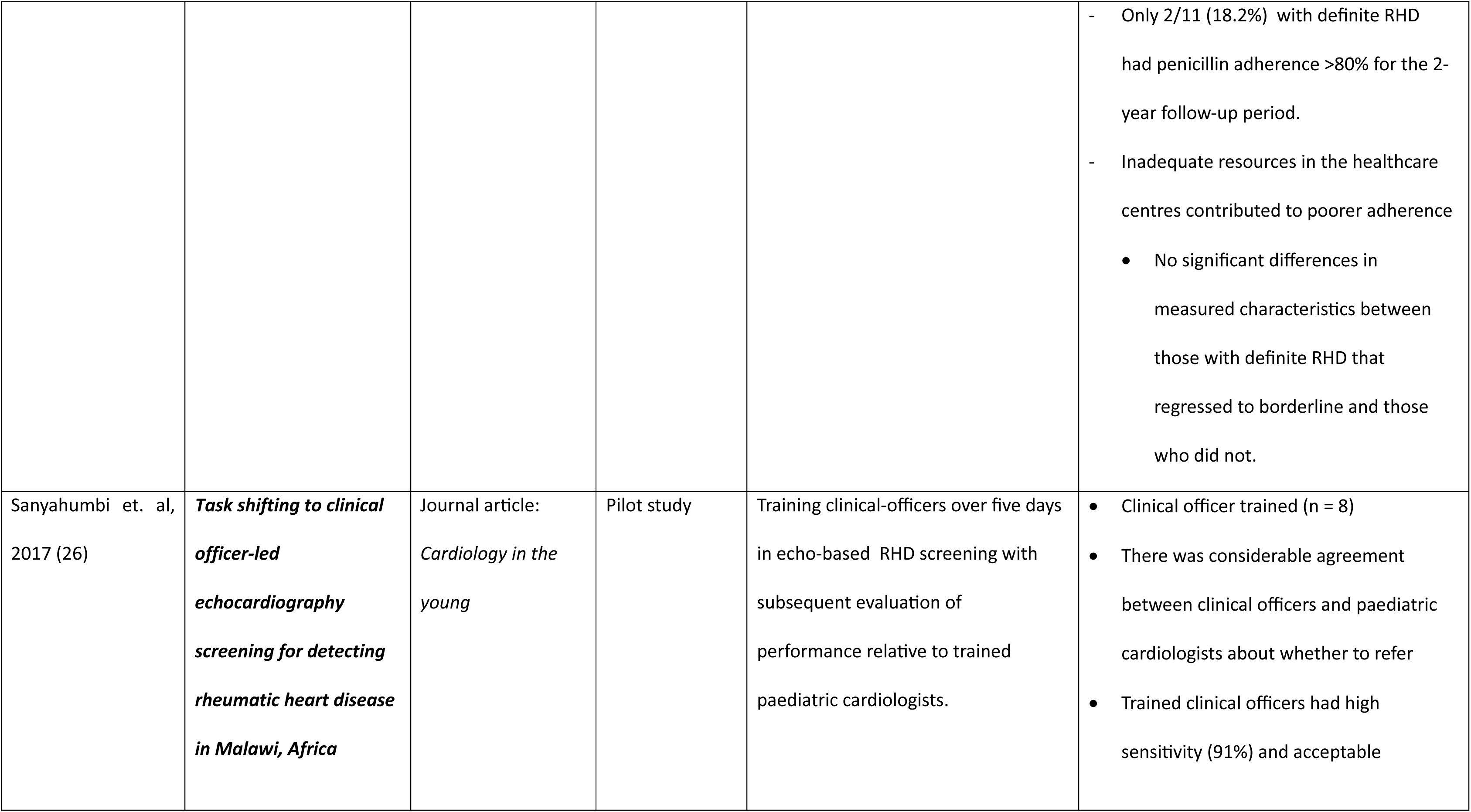
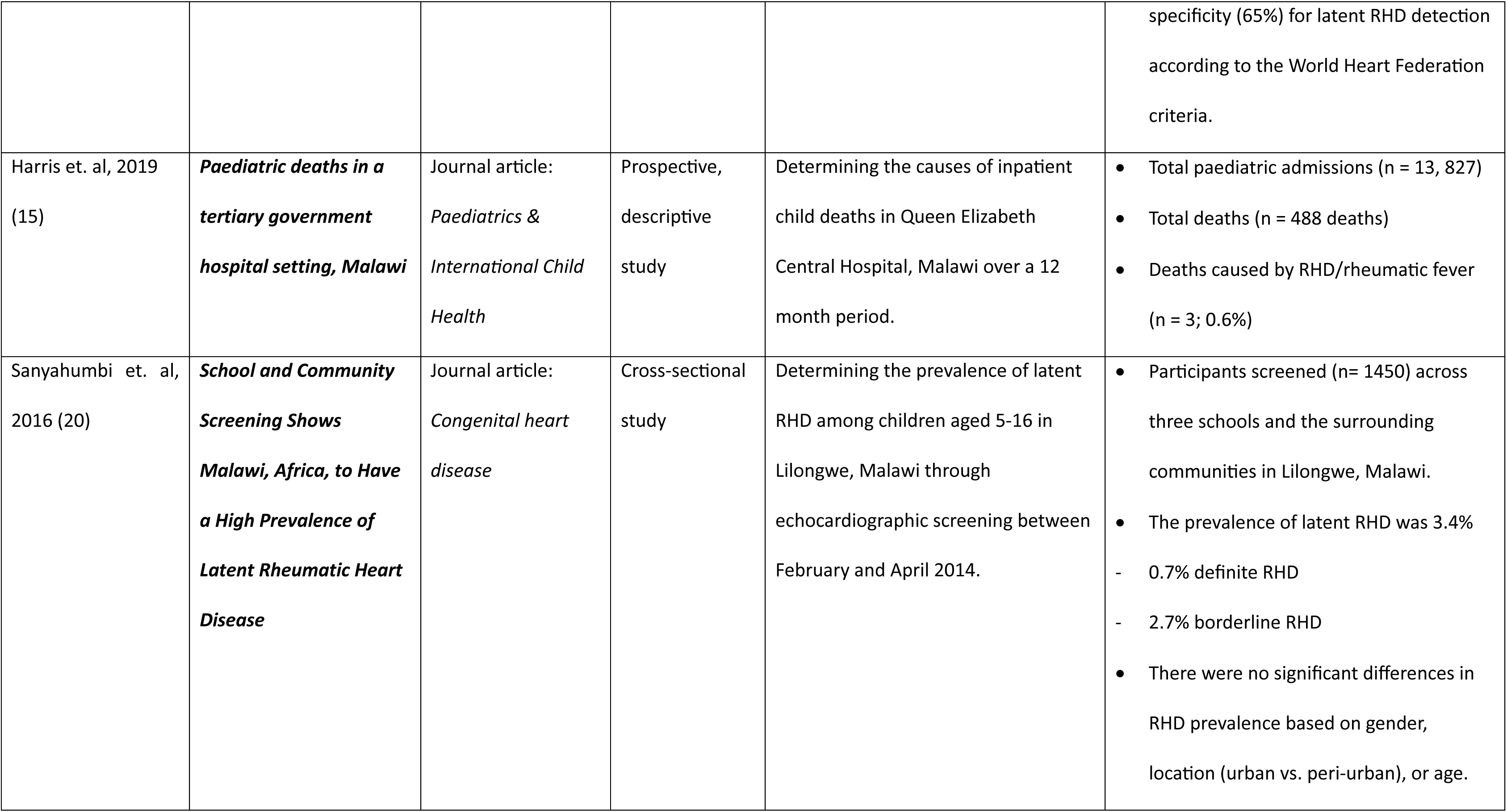
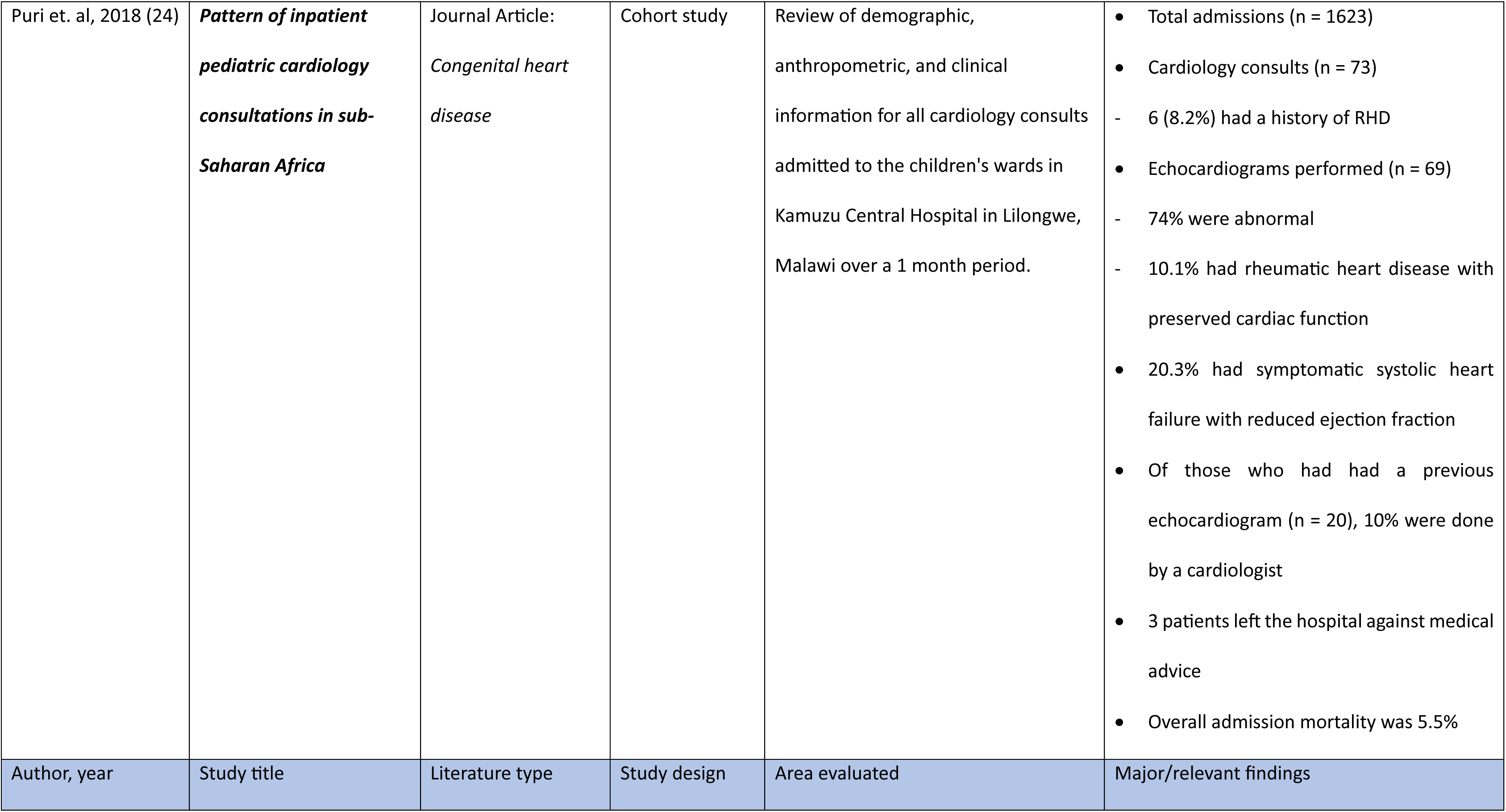
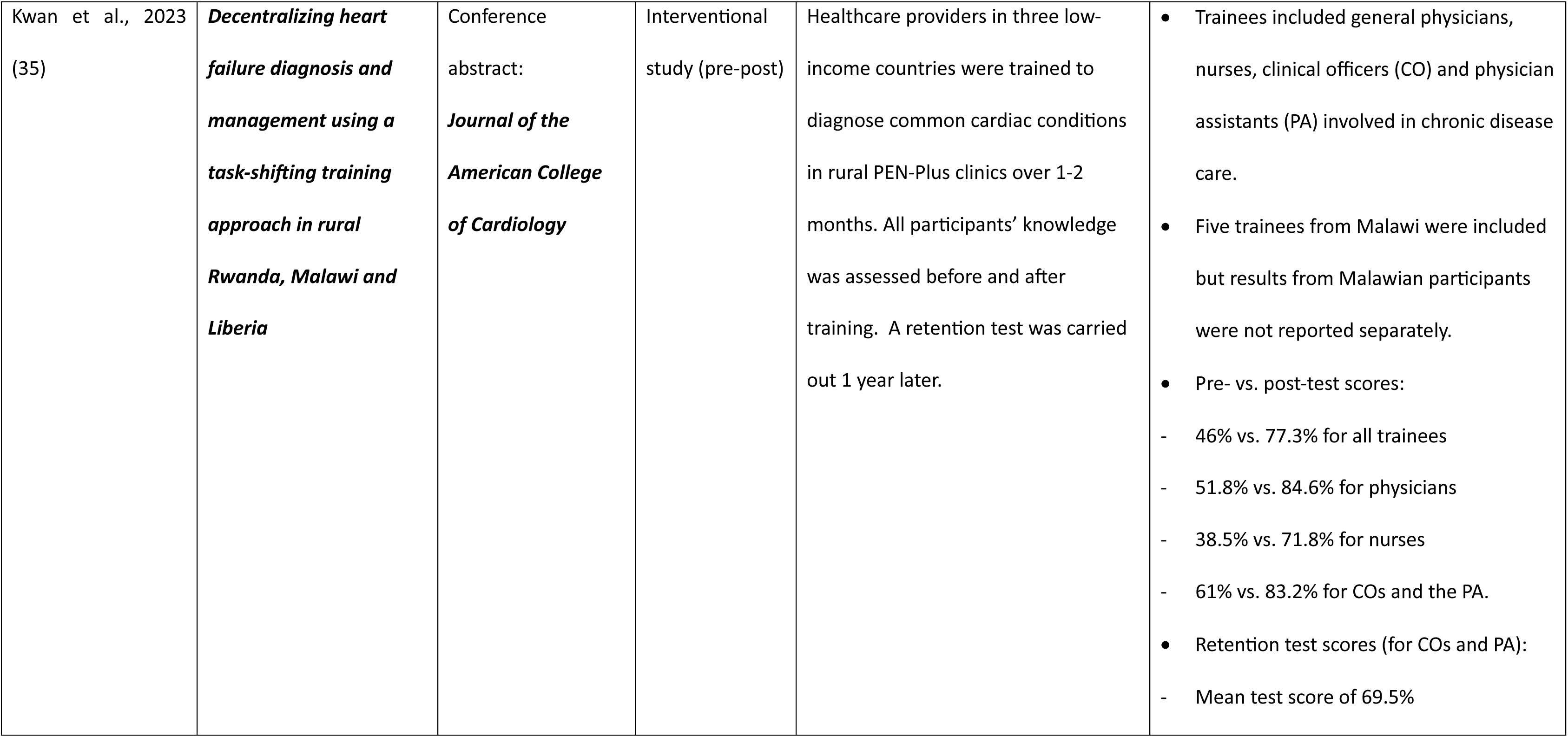
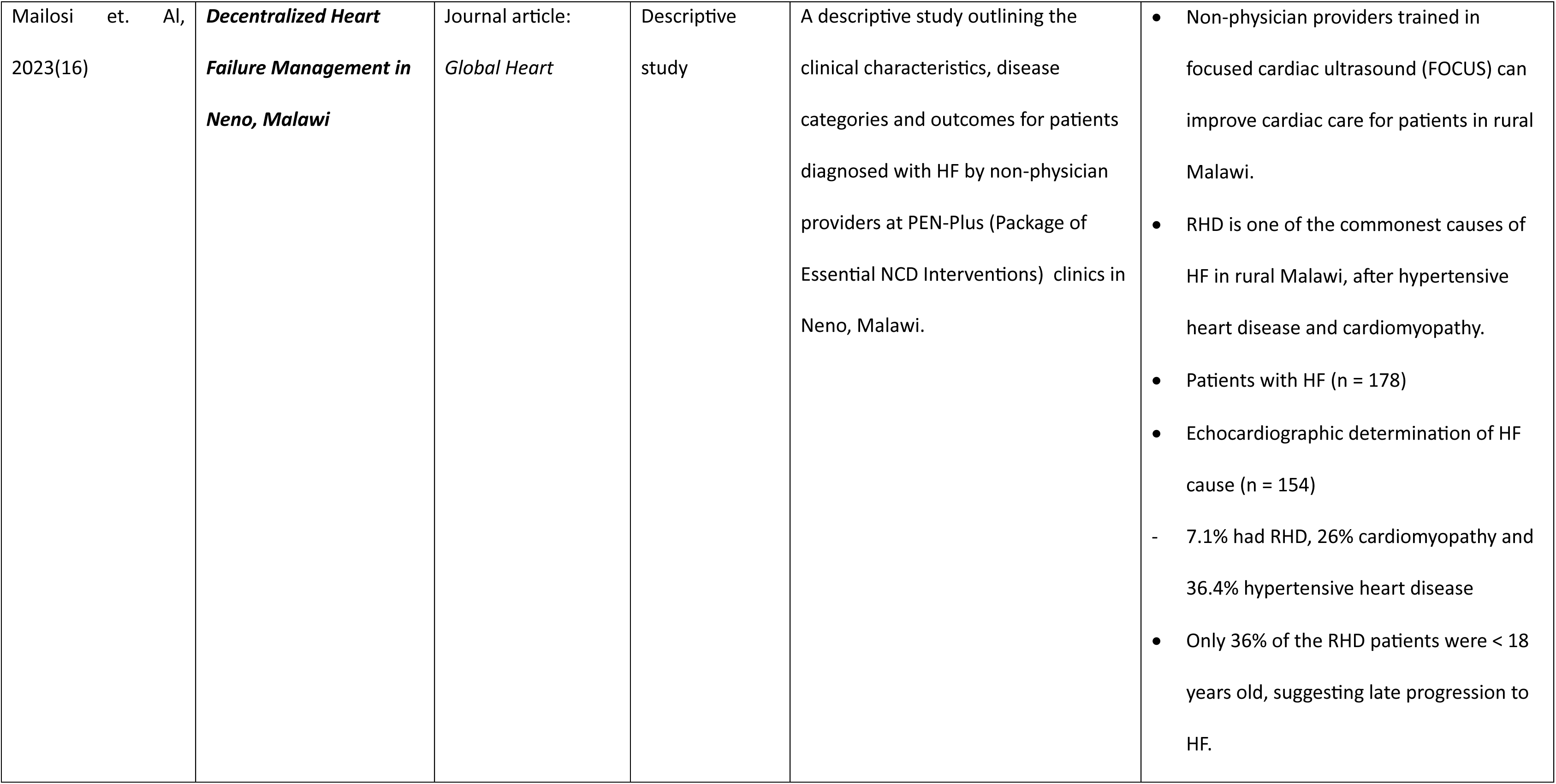
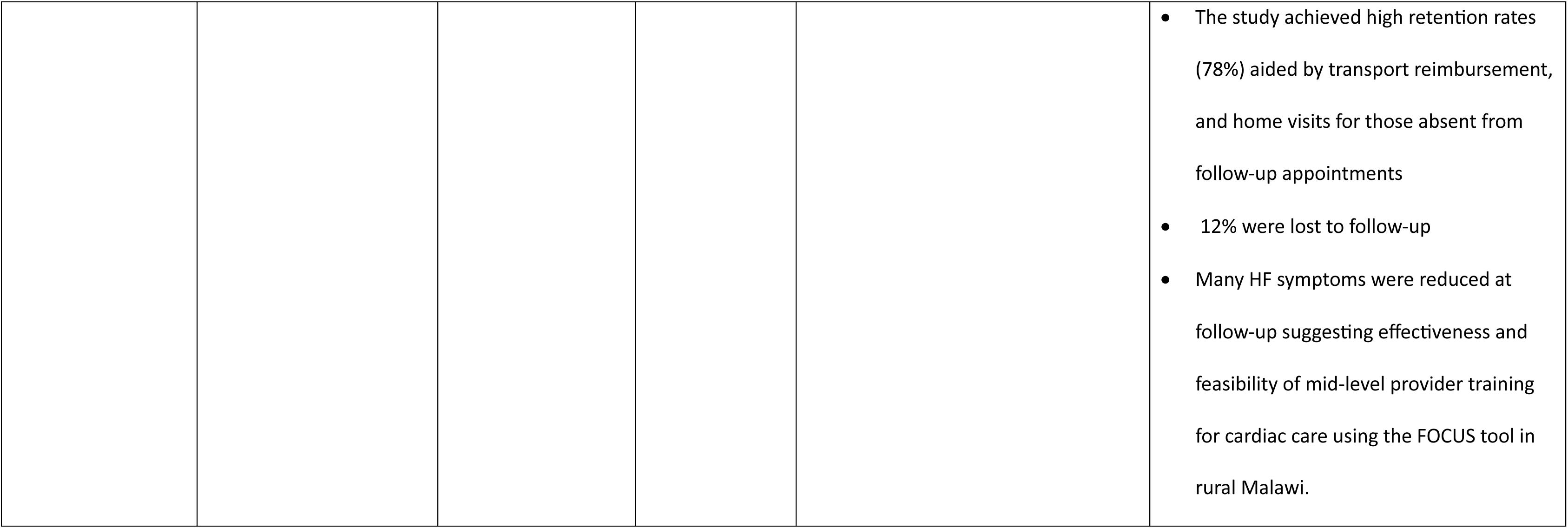
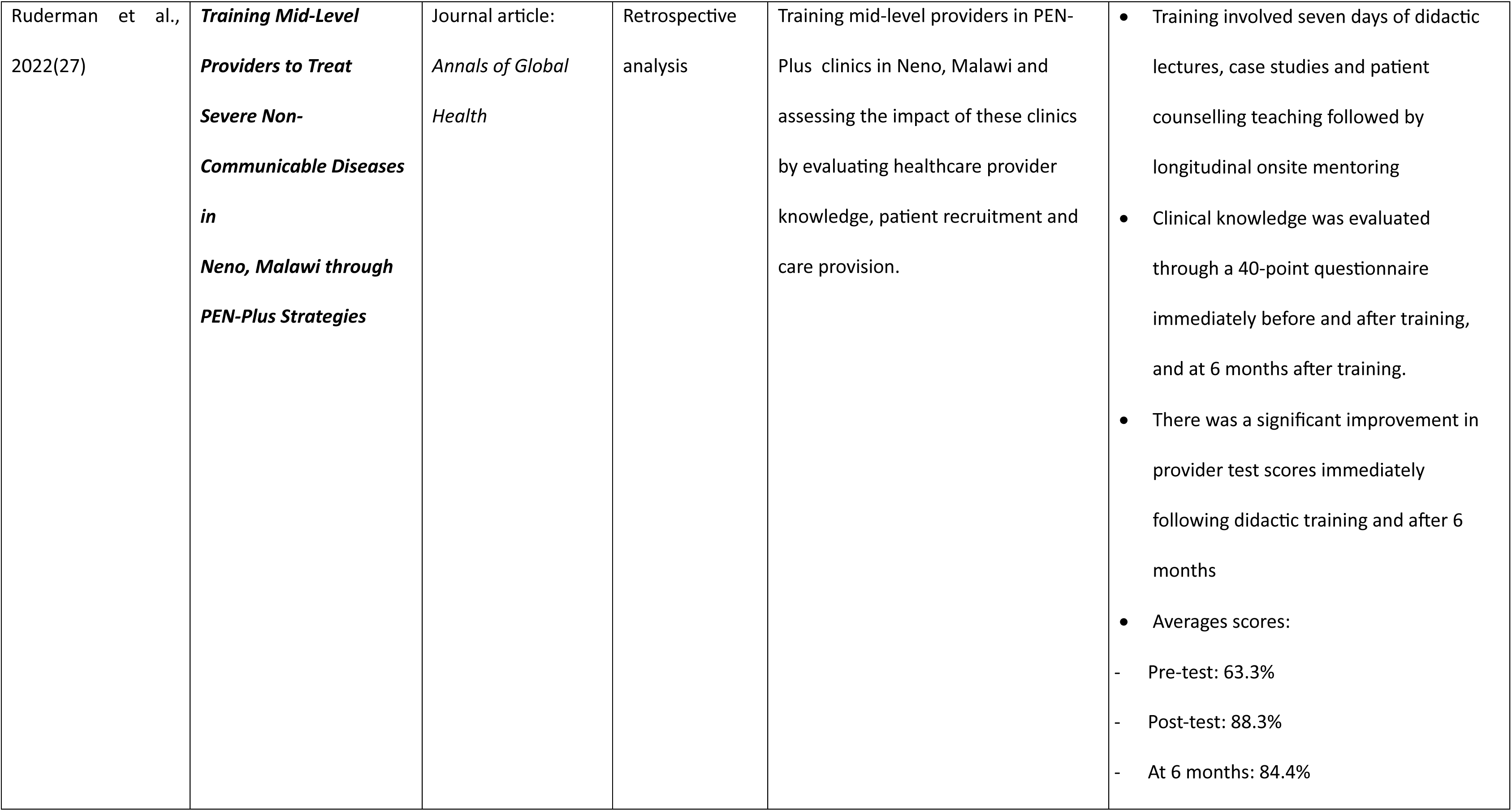
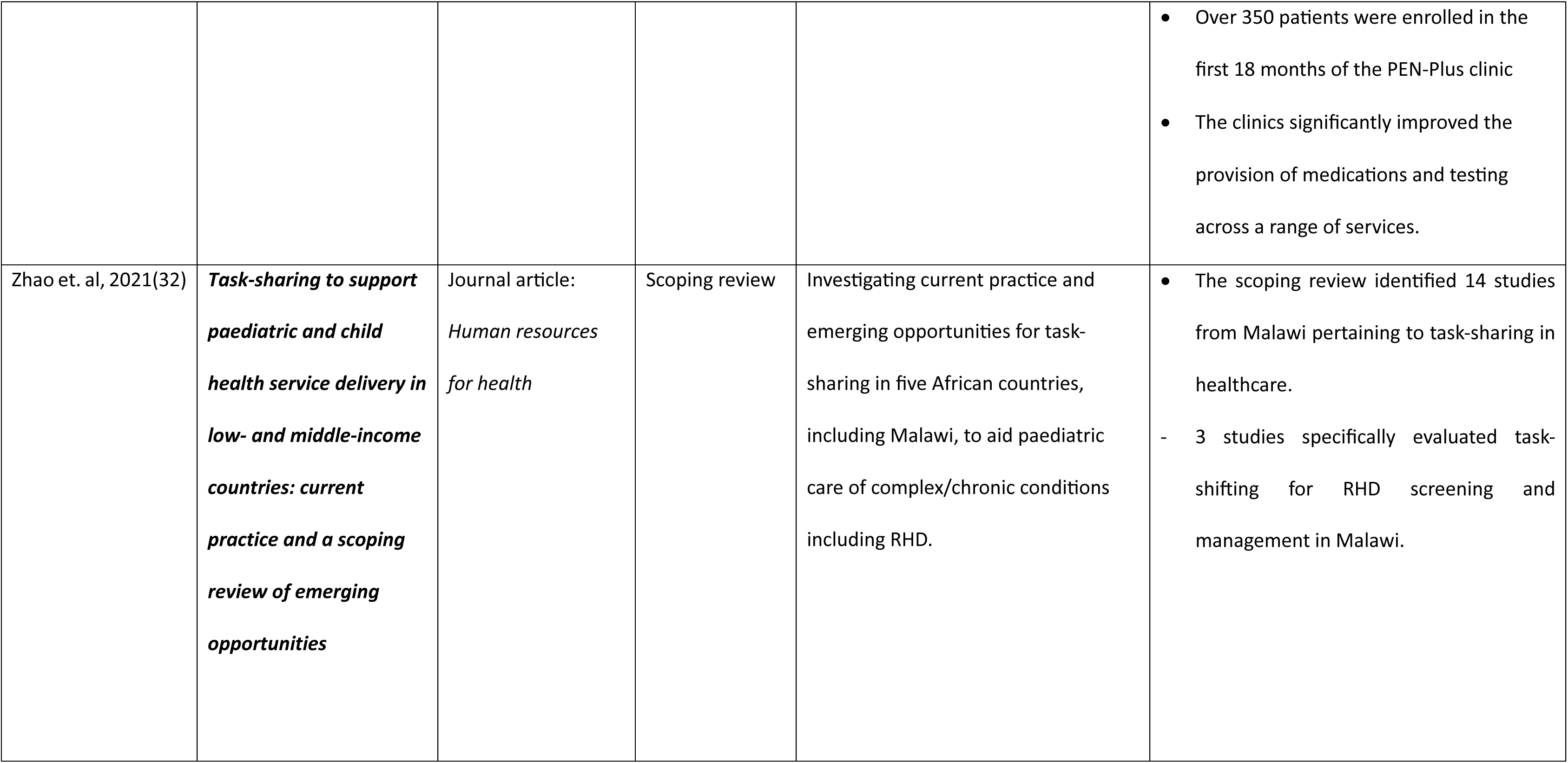
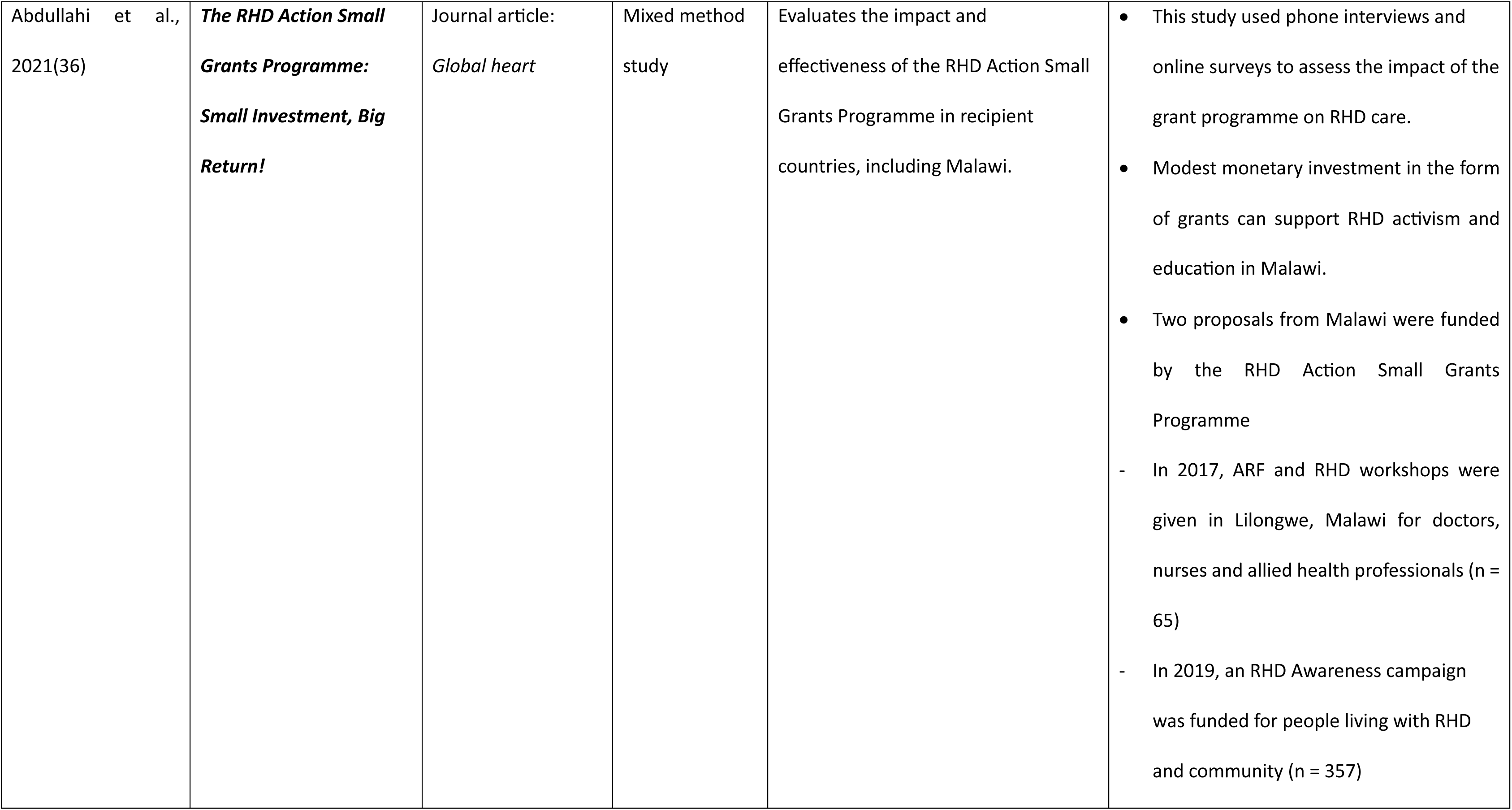
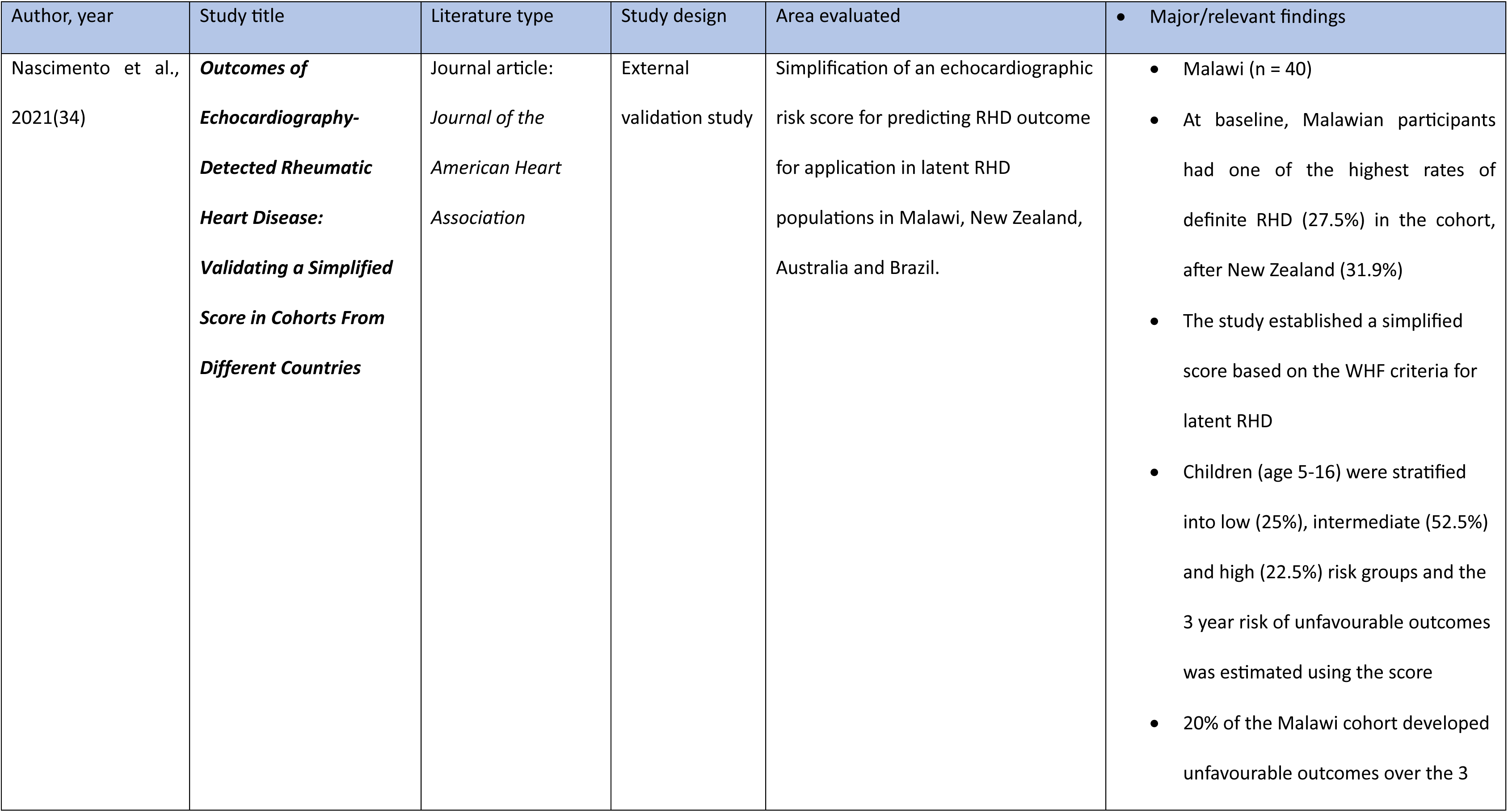
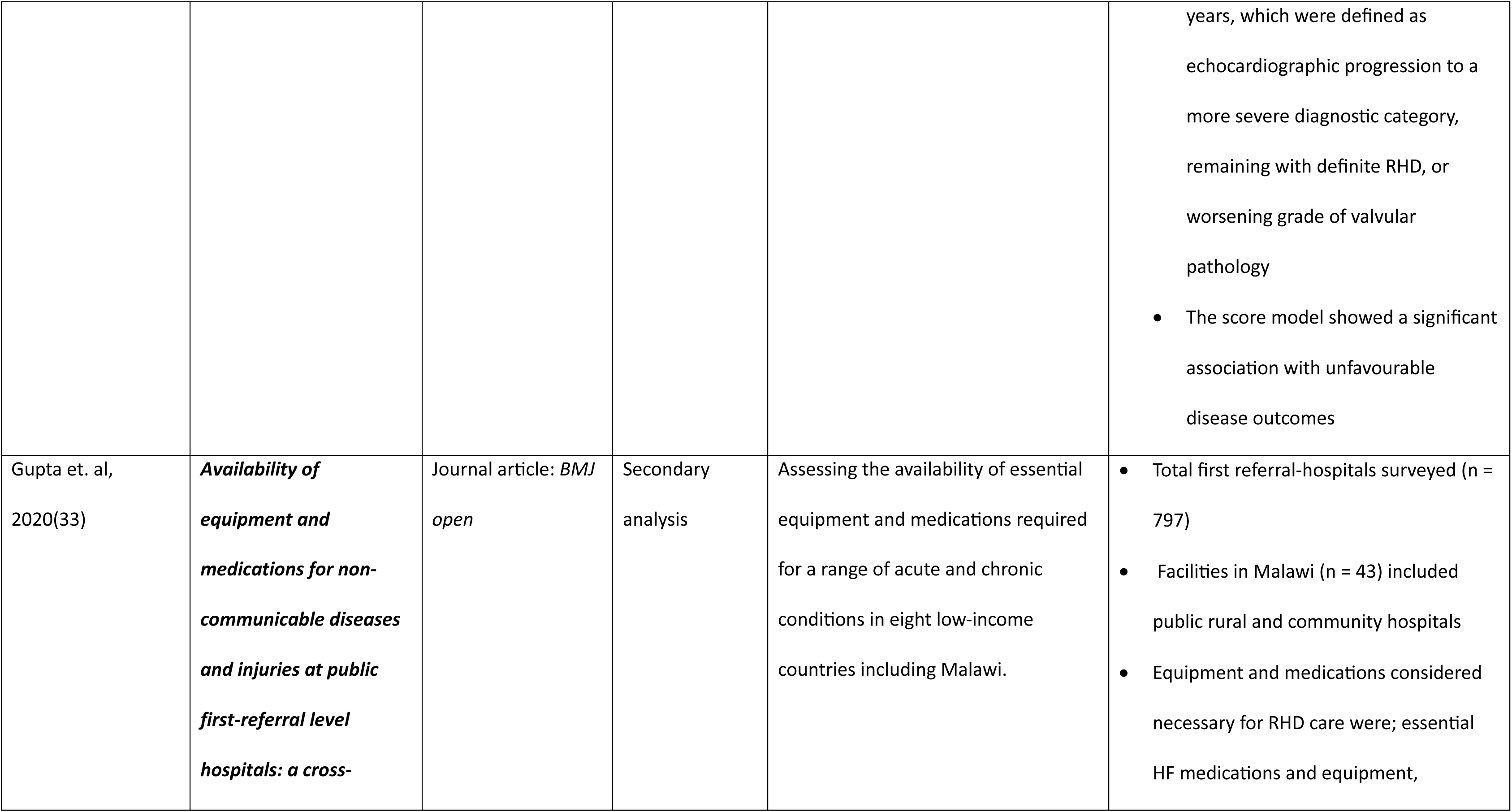
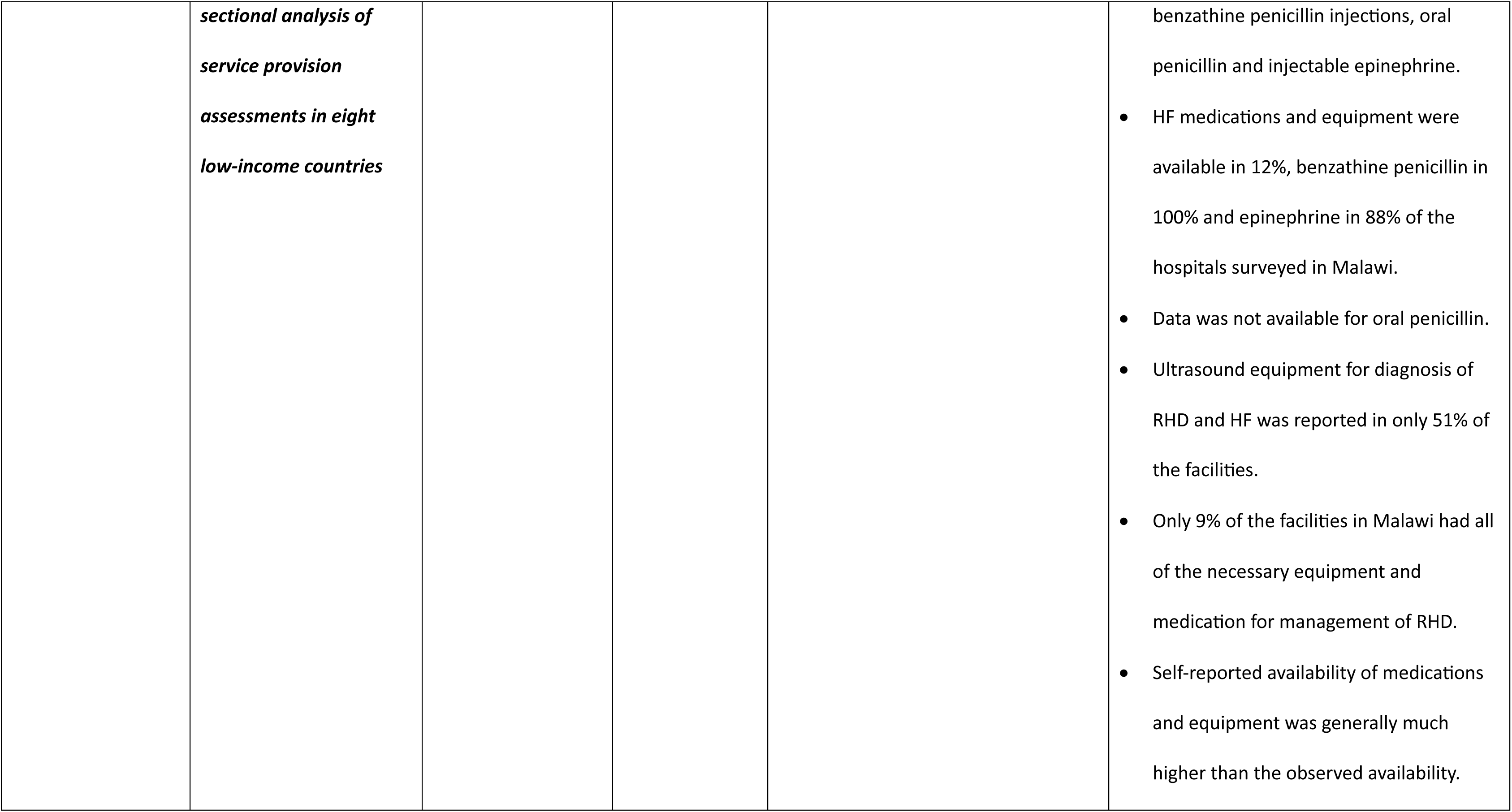
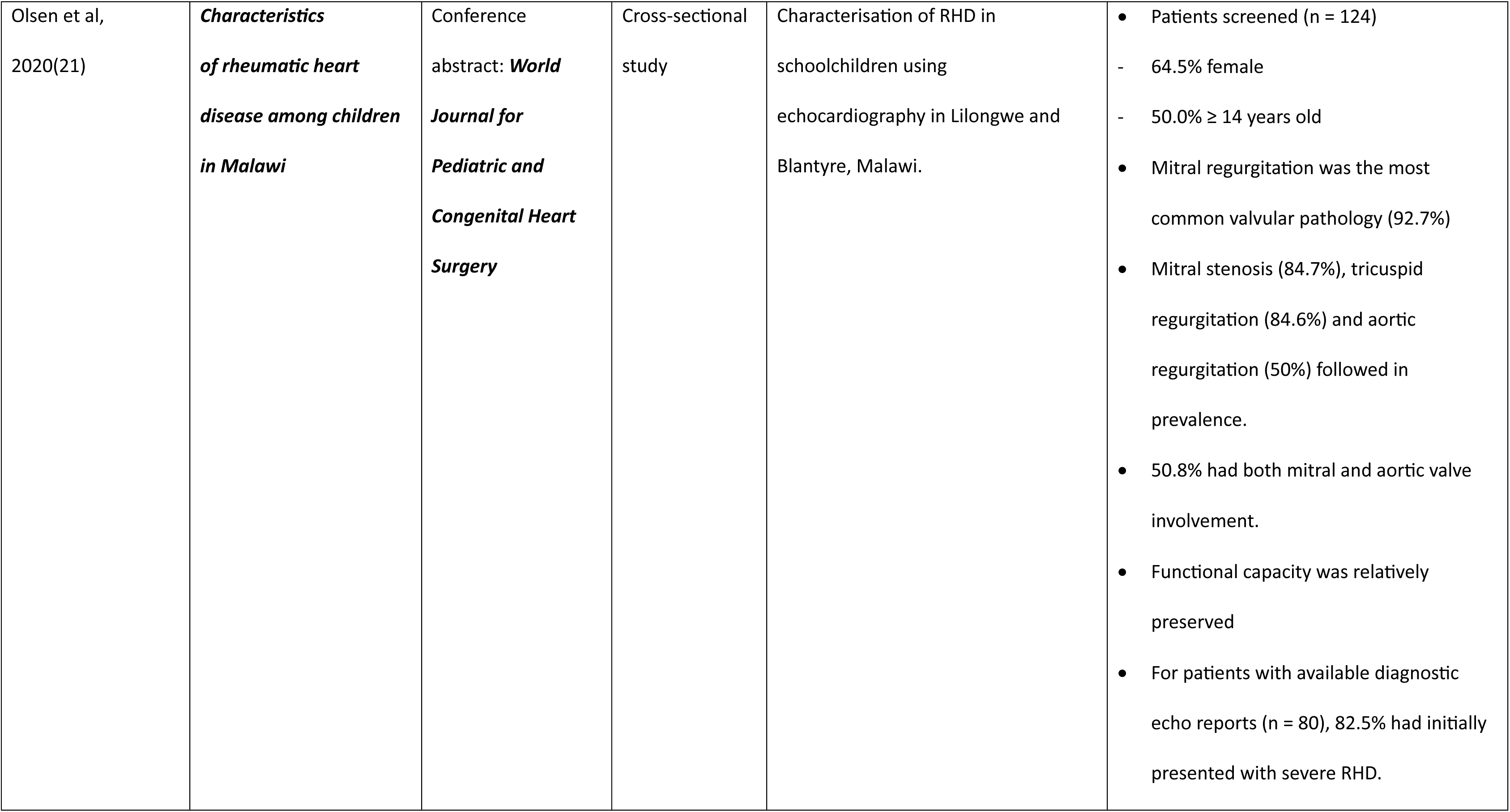
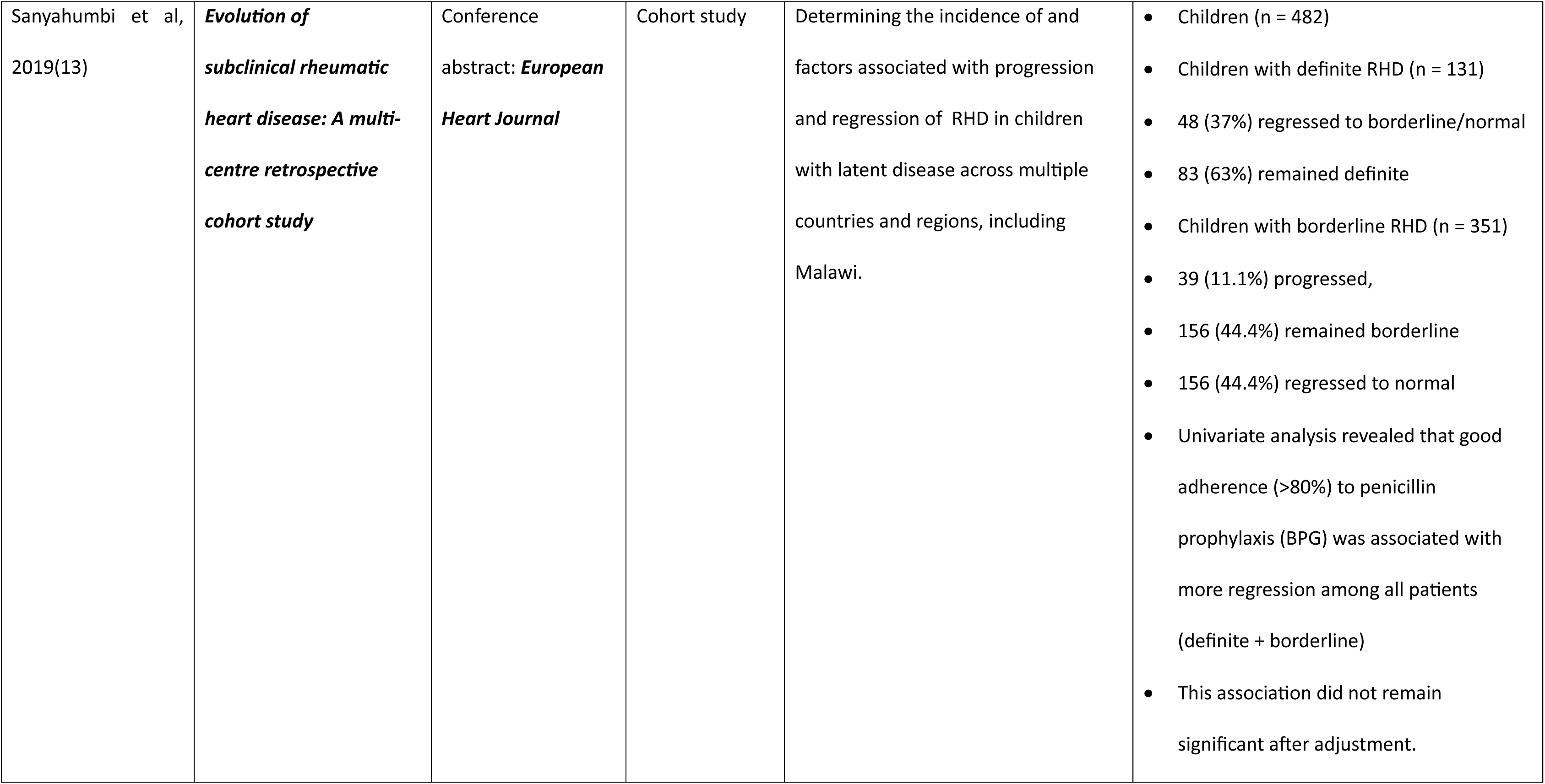
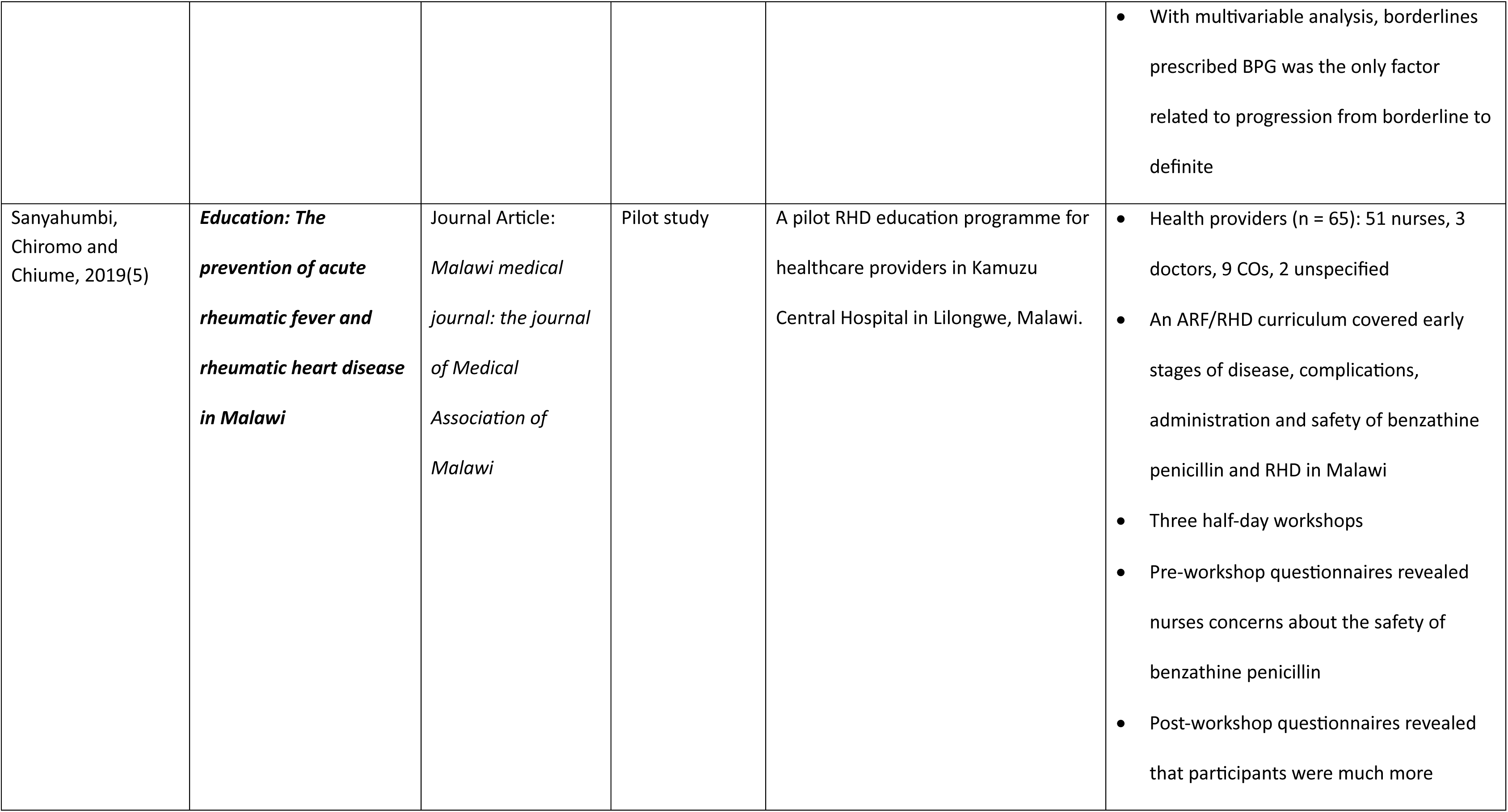
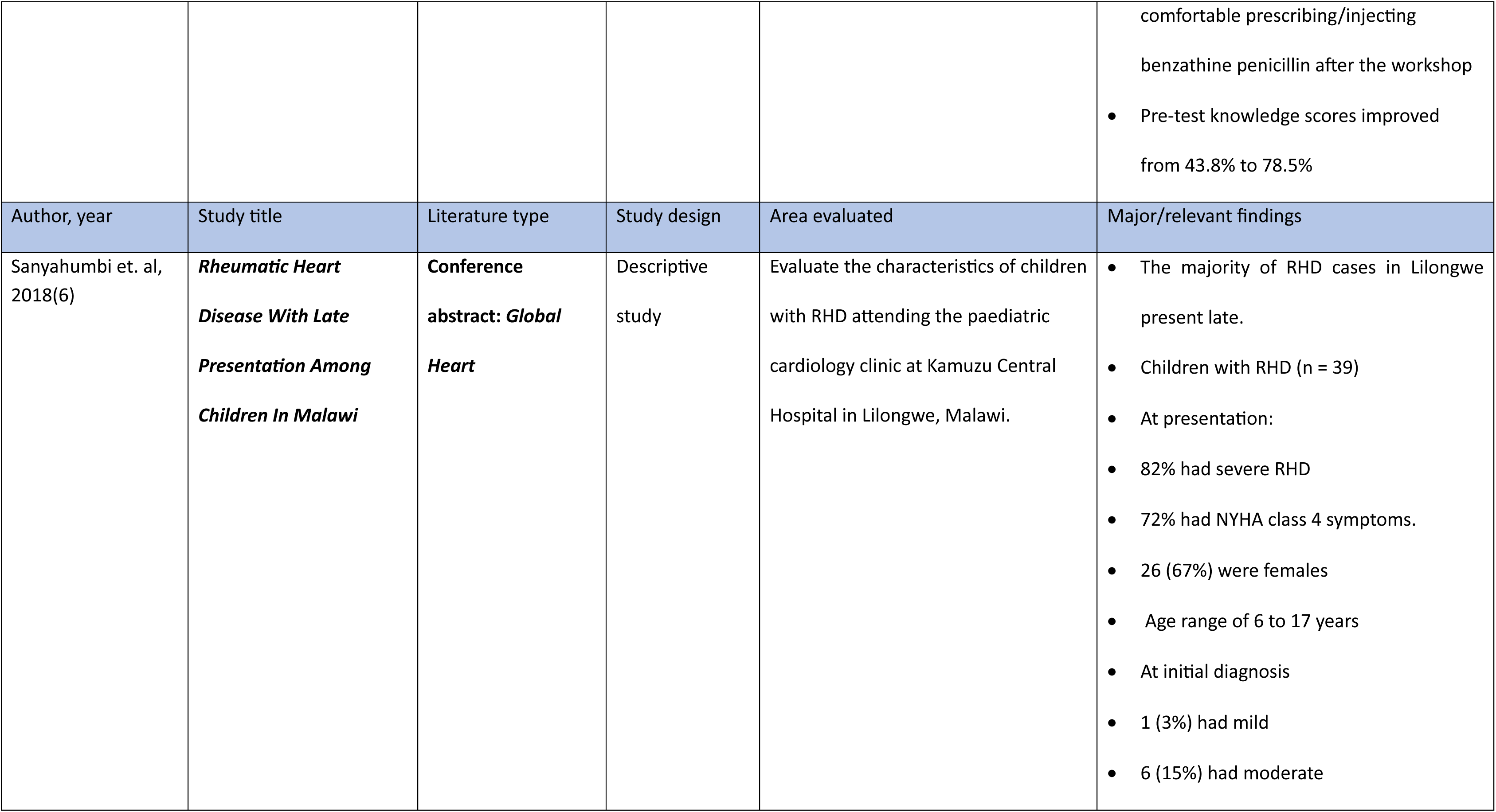
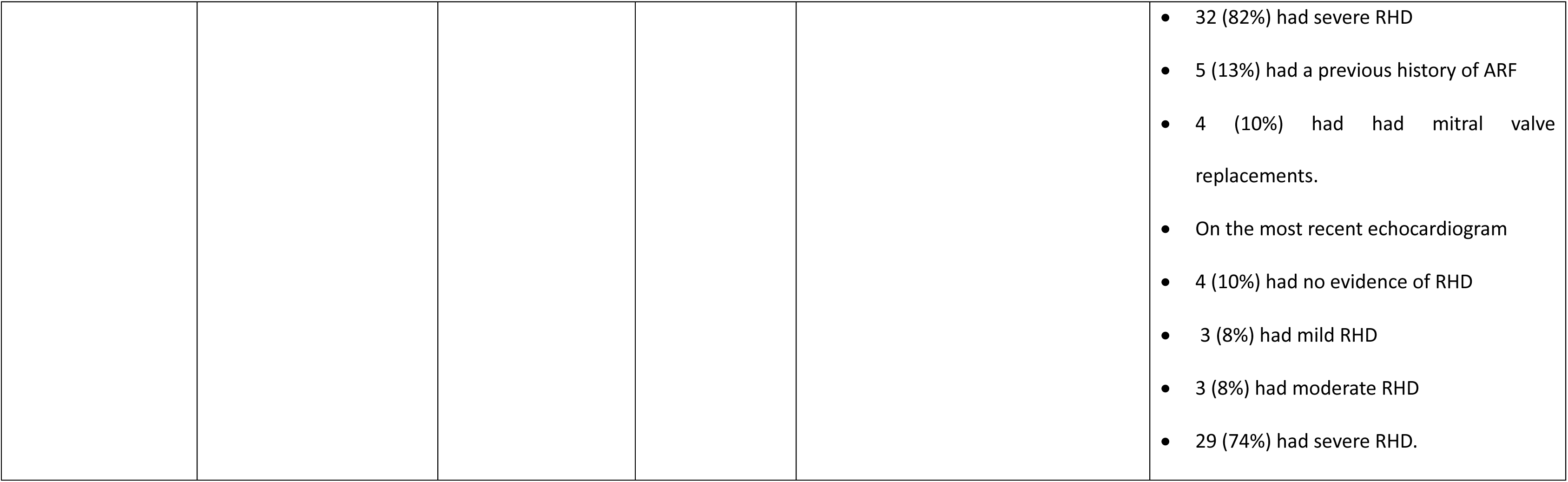
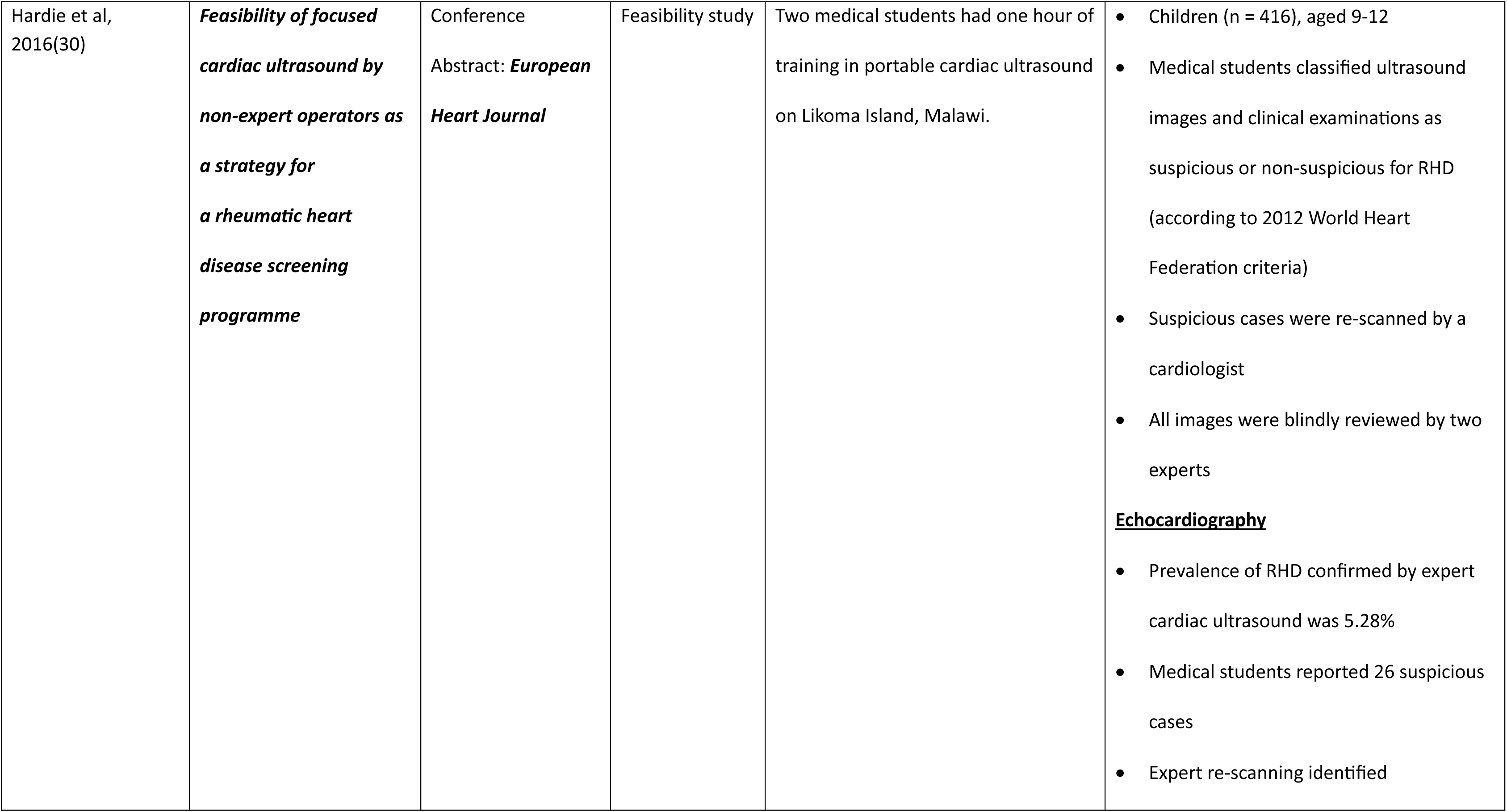
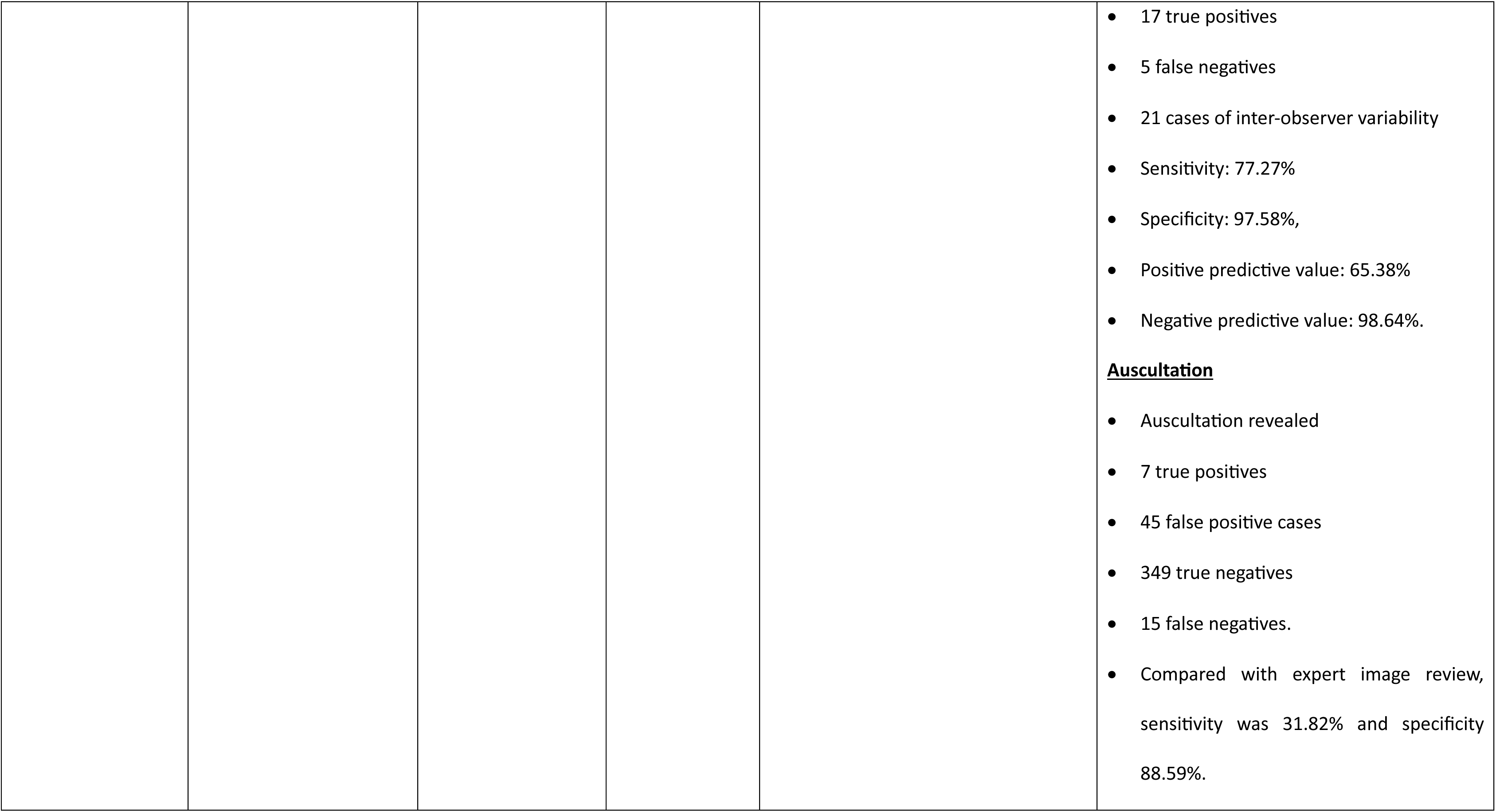
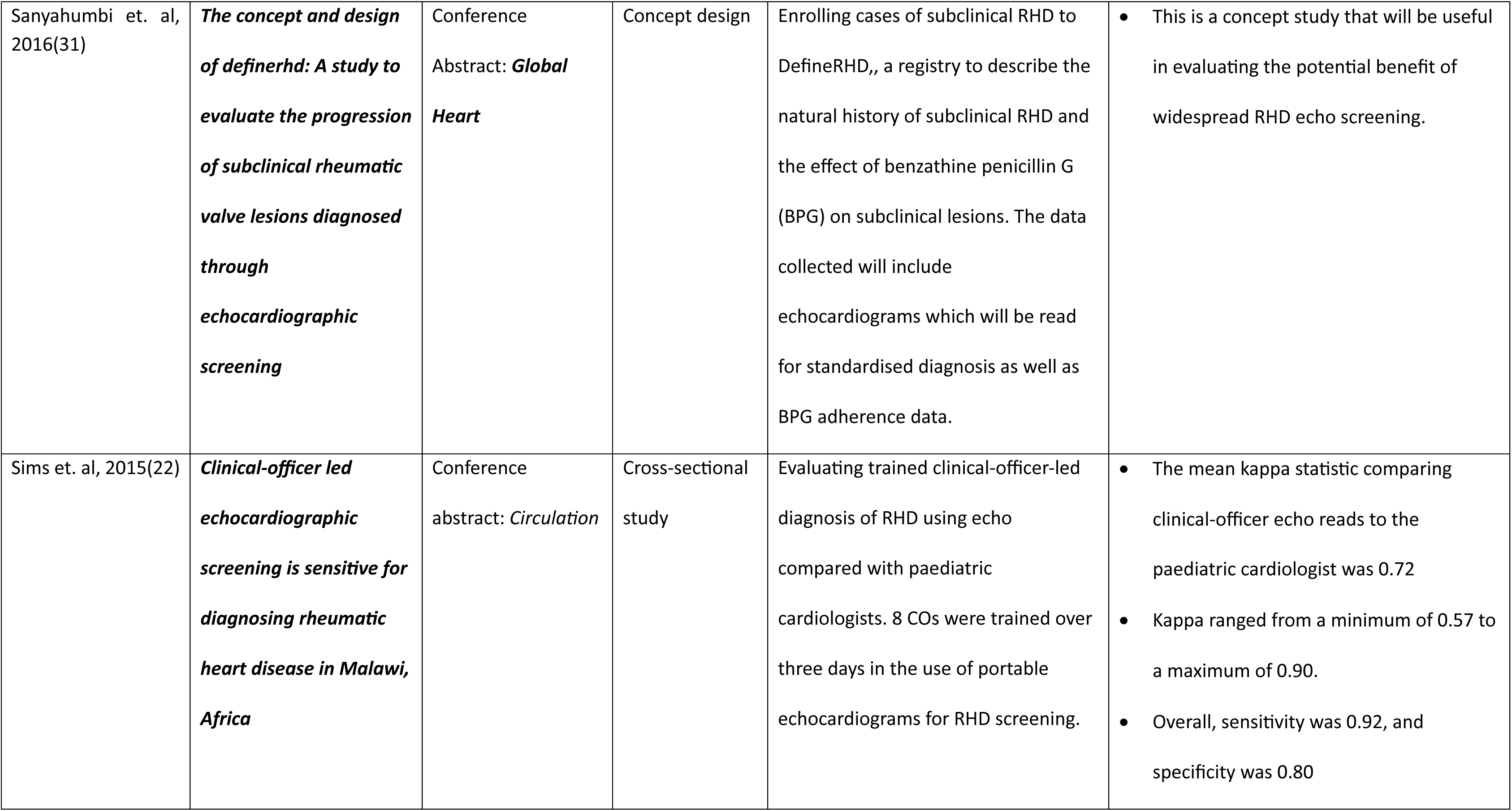
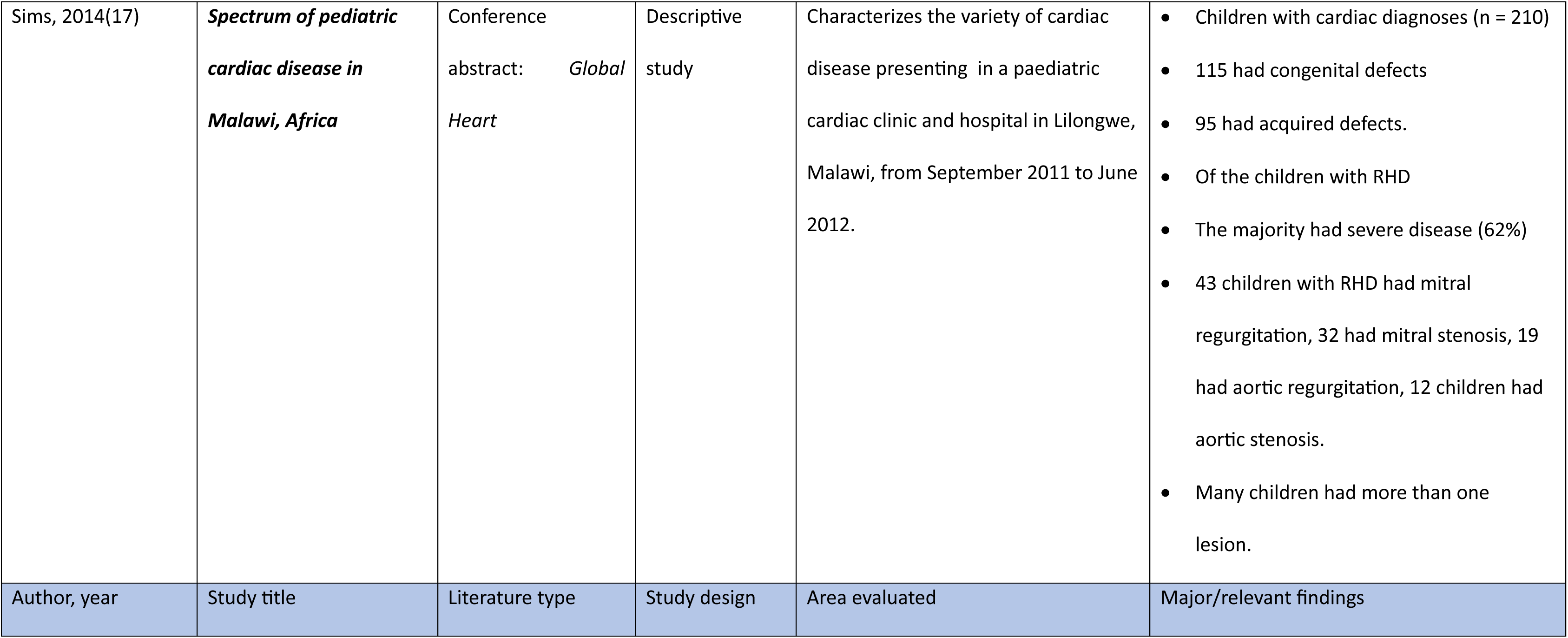
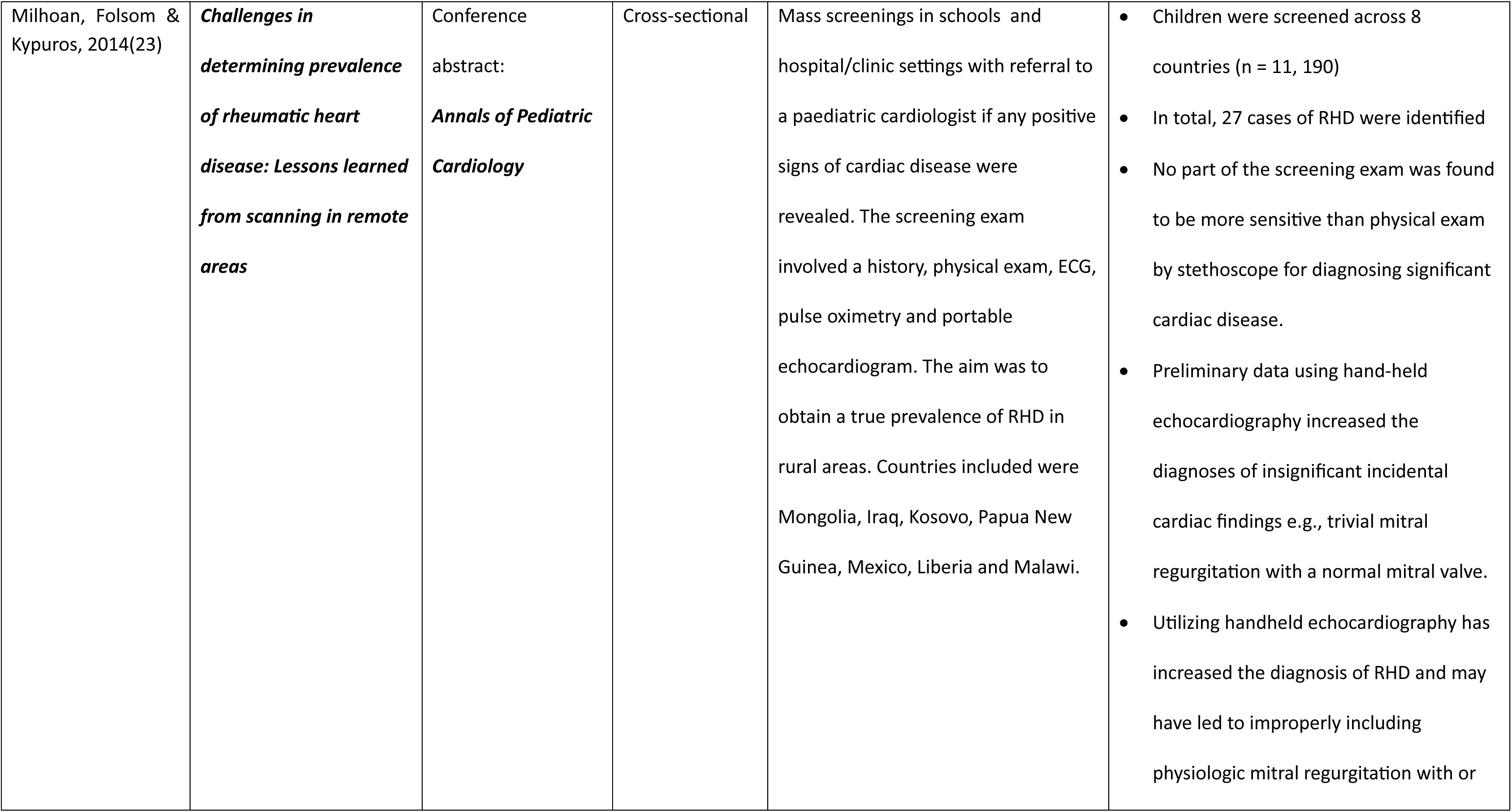
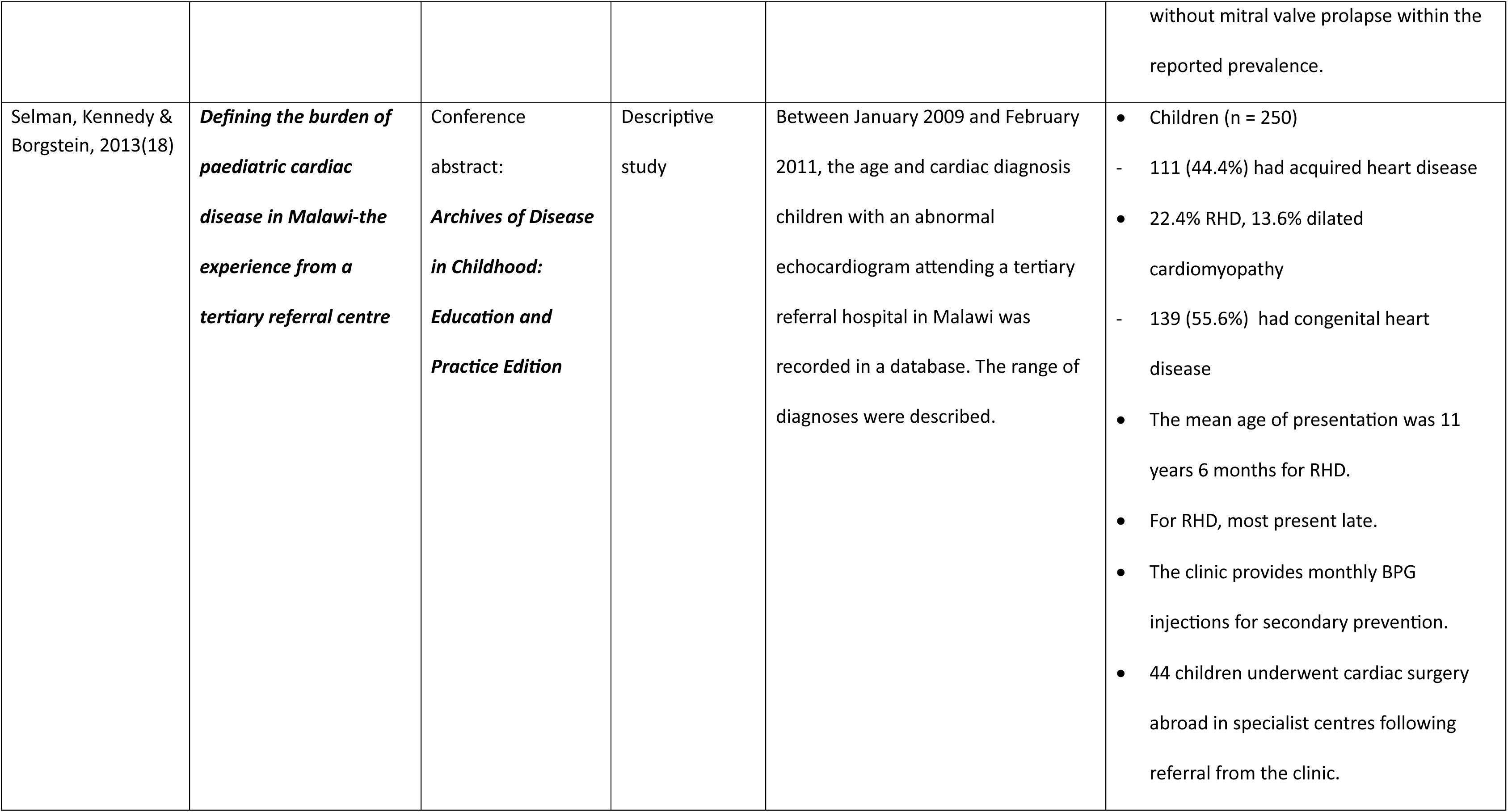
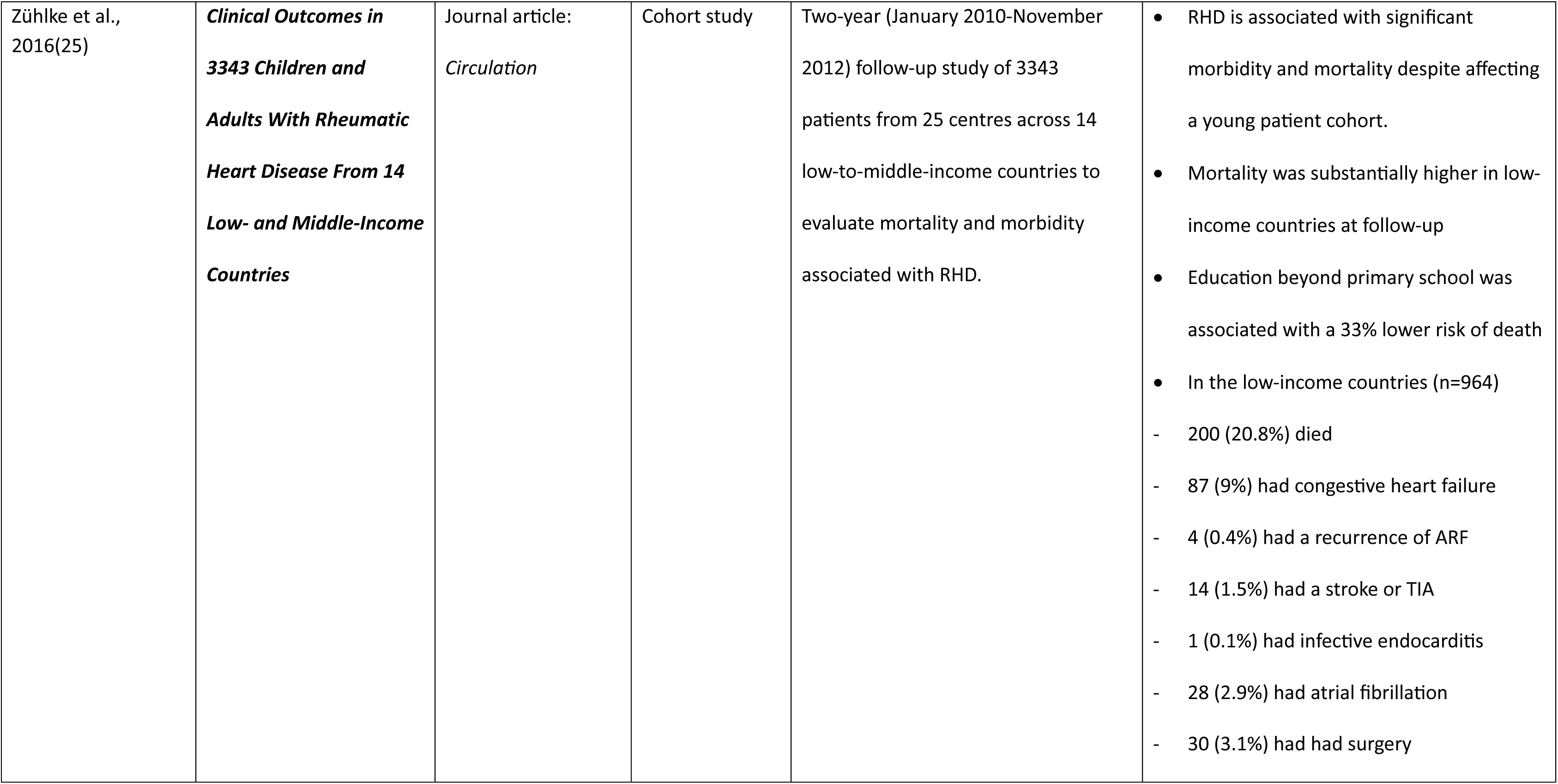
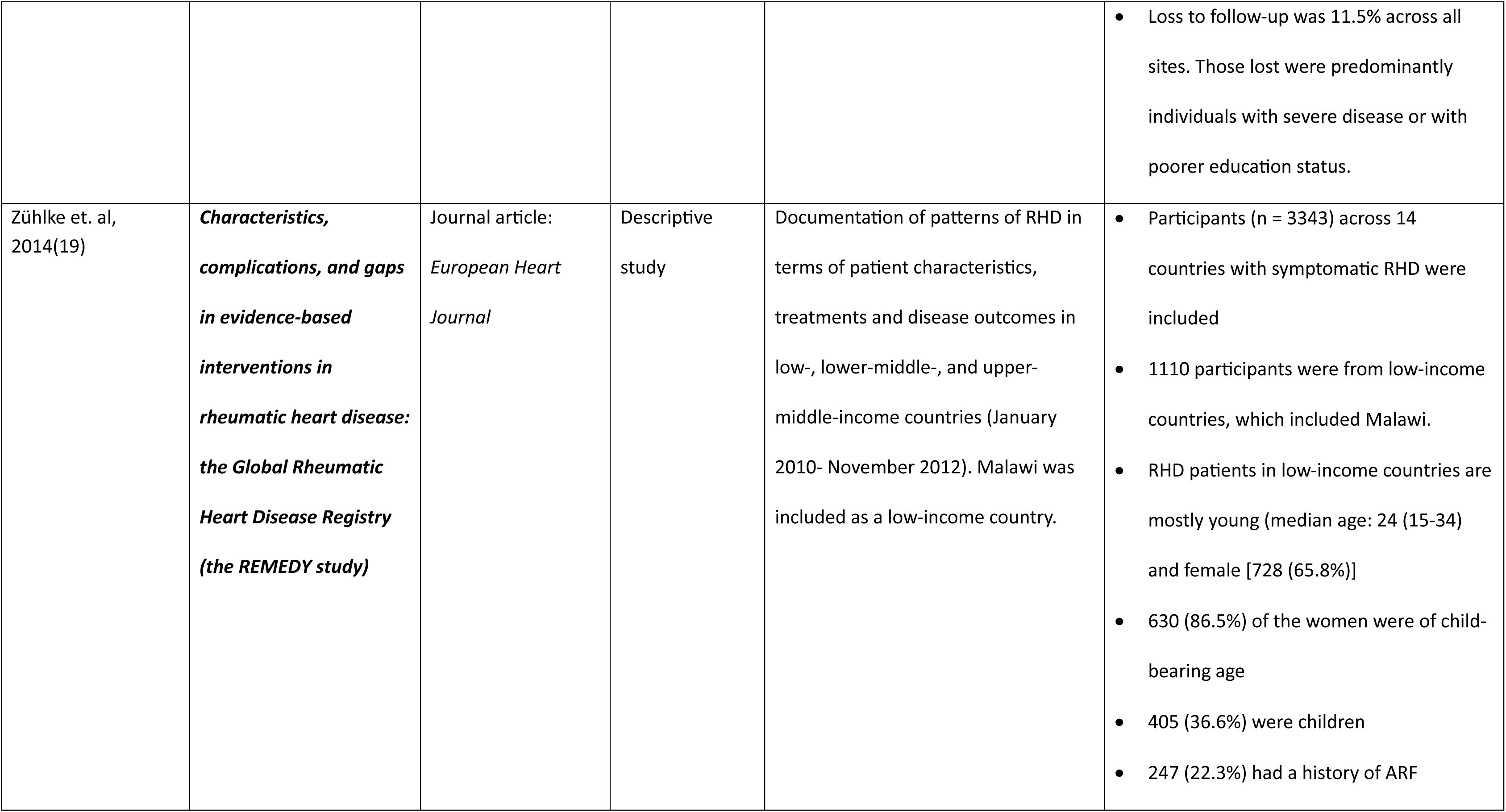
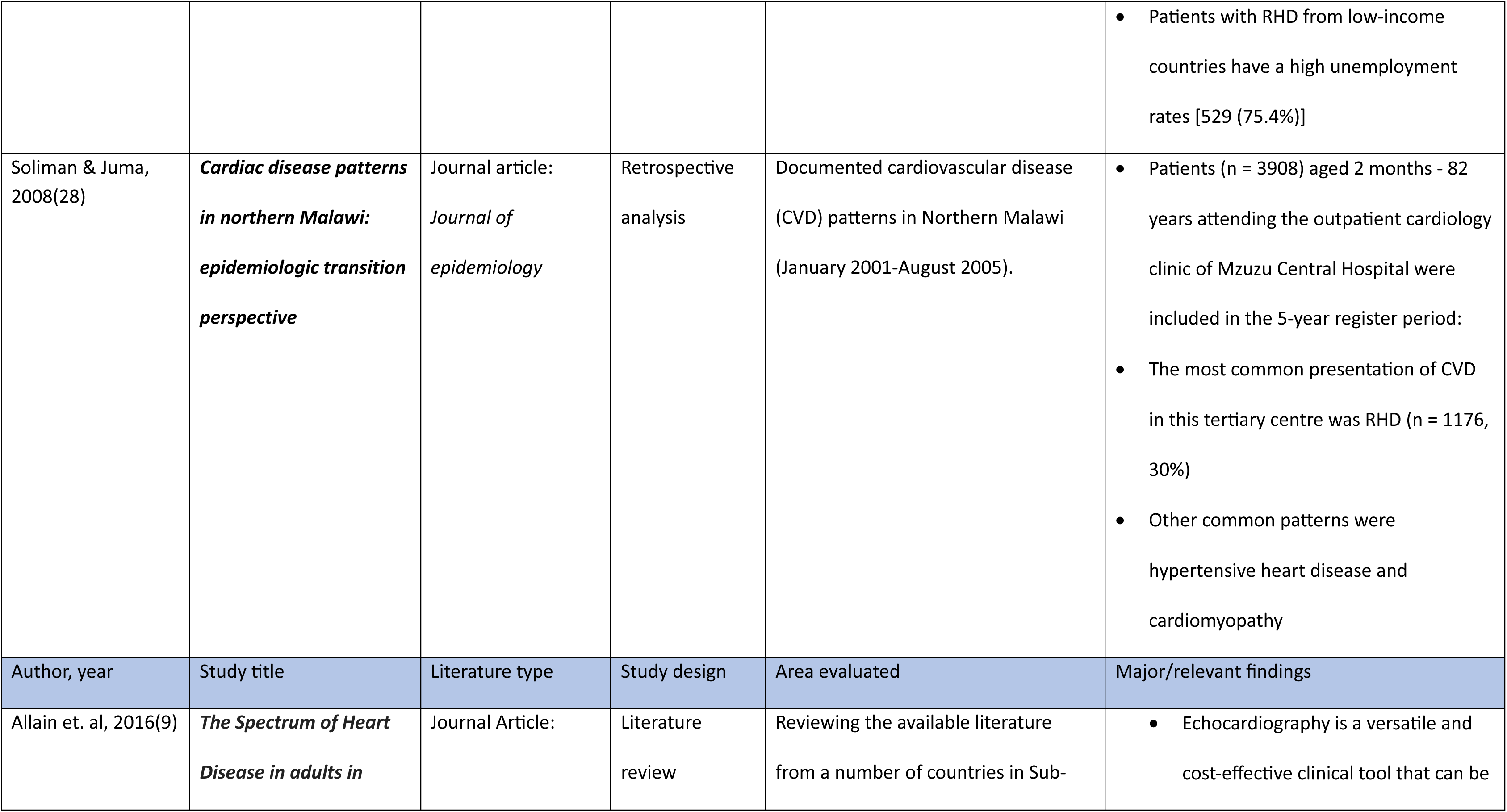
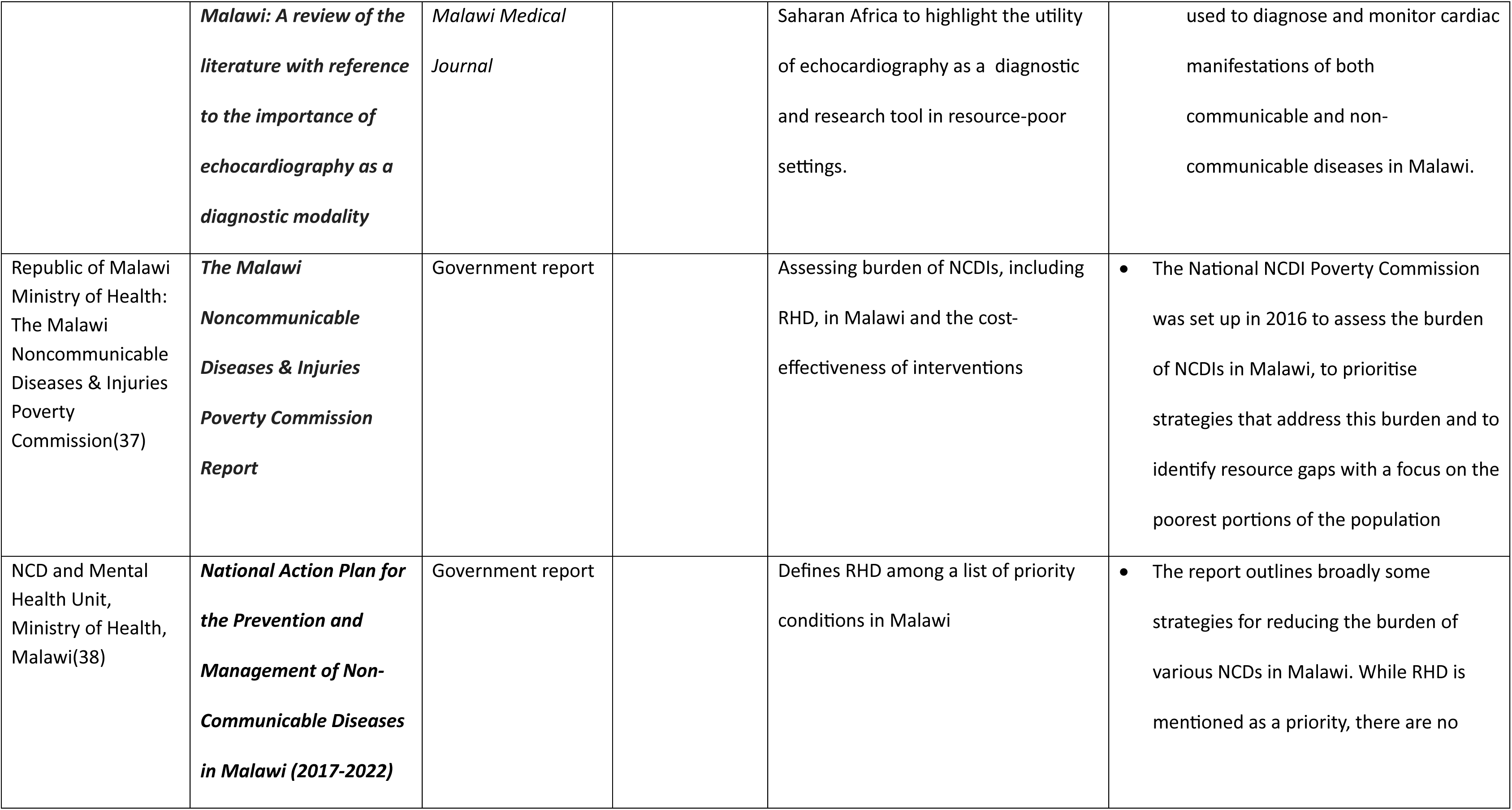
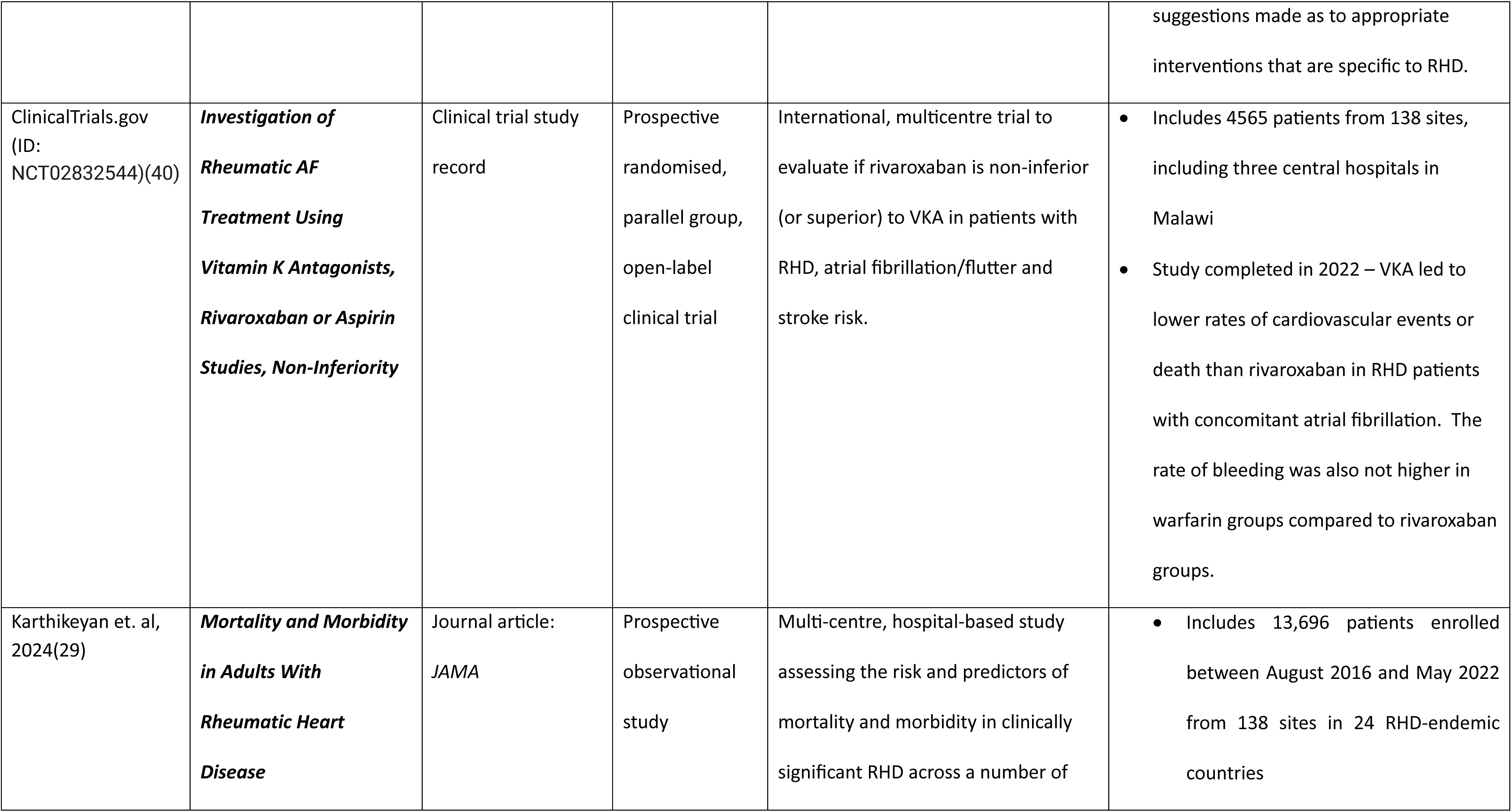
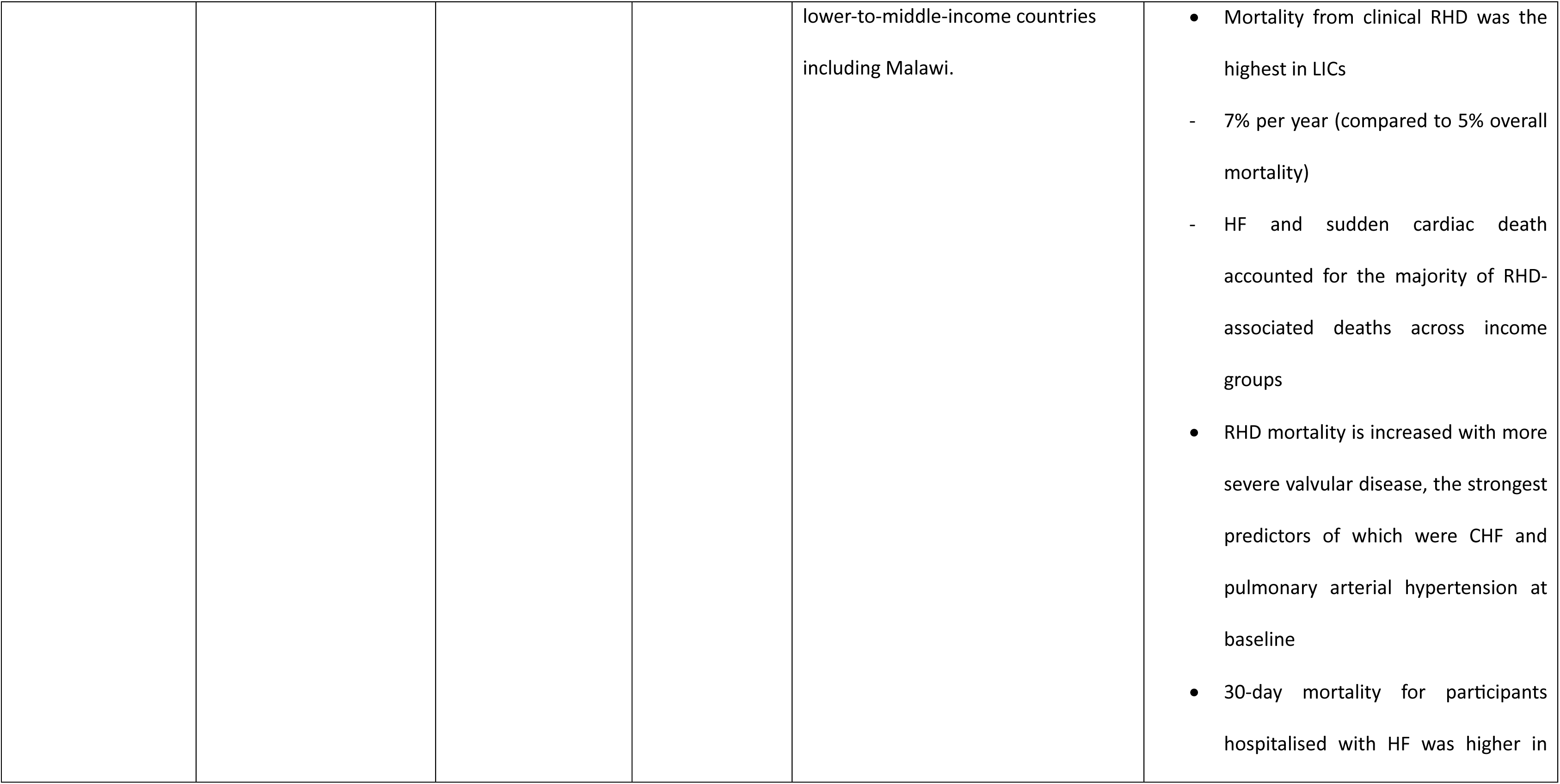
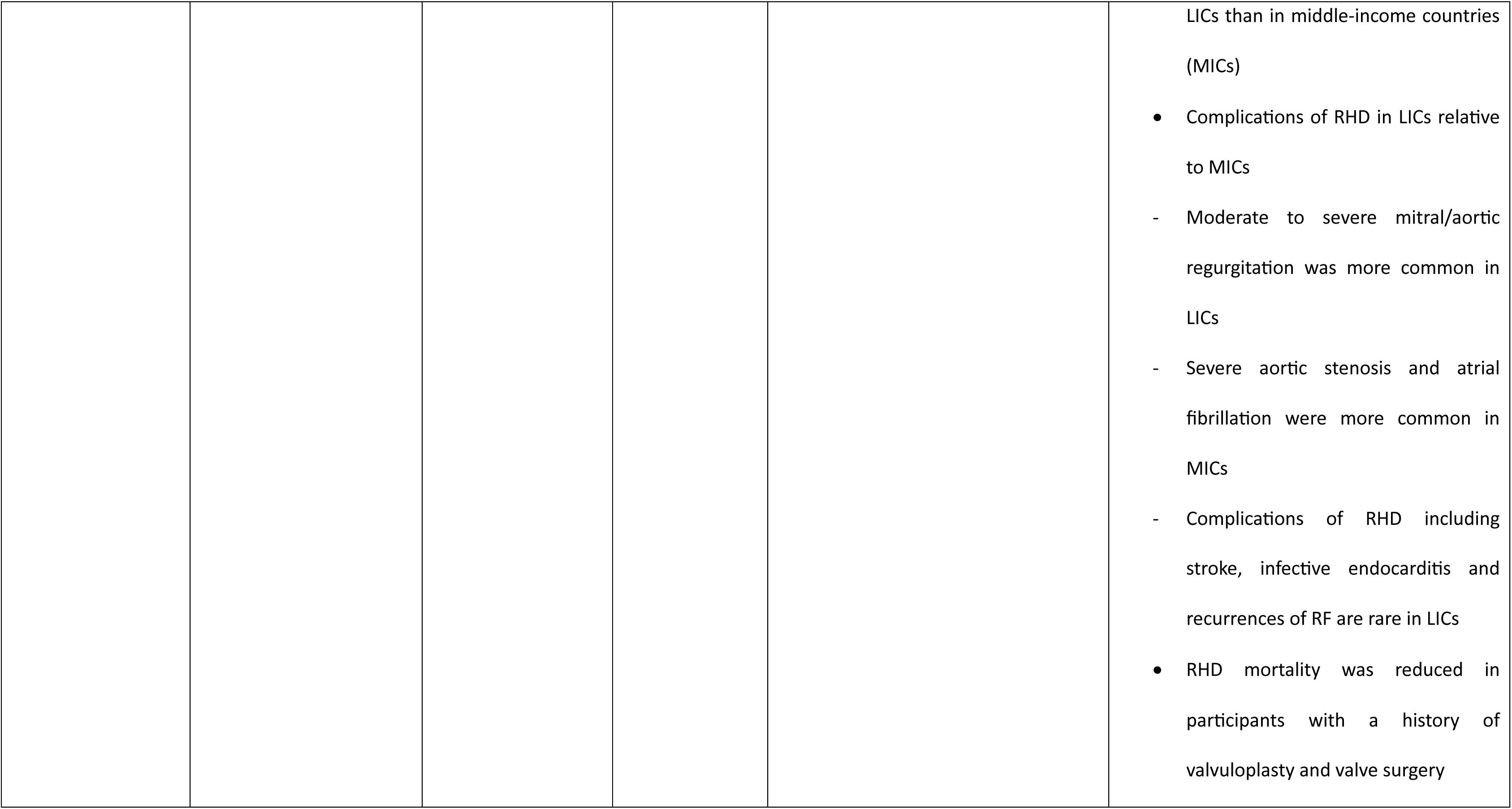
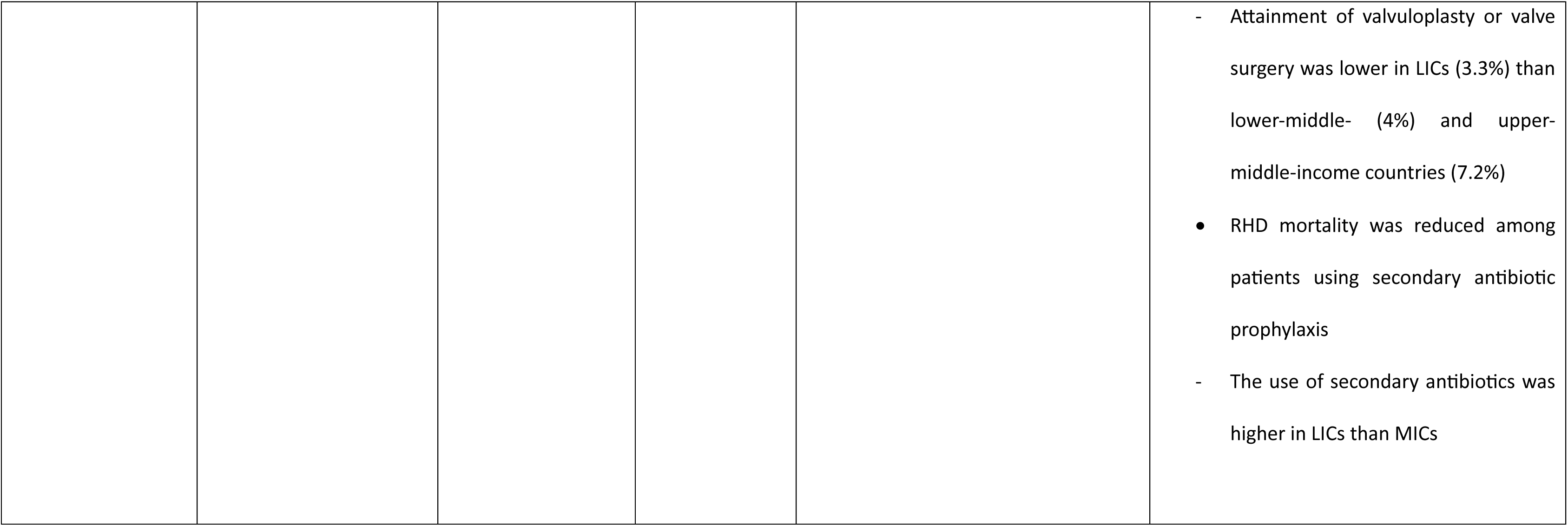
DescripIon of studies included.

## S2. Search strategies

### PubMed search strategy: performed 24/08/2024

(“Rheumatic Heart Disease”[MeSH Terms] OR “rheumatic heart disease*”[Title/Abstract] OR “rheumatic heart”[Title/Abstract] OR “rheumatic carditis”[Title/Abstract] OR

(“Rheumatic Fever”[MeSH Terms] OR “rheumatic fever*”[Title/Abstract] OR “acute rheumatic fever*”[Title/Abstract]) OR

(“Streptococcus pyogenes”[MeSH Terms:noexp] OR “Pharyngitis”[MeSH Terms] OR “Group A streptococcus”[Title/Abstract] OR “group a streptococcal infection*”[Title/Abstract]))

AND

(“Cost of Illness”[MeSH Terms] OR “Morbidity”[MeSH Terms] OR “Mortality”[MeSH Terms] OR “Prevalence”[MeSH Terms] OR “Incidence”[MeSH Terms] OR

((“Diagnosis”[MeSH Terms] OR “Referral and Consultation”[MeSH Terms] OR “Therapeutics”[MeSH Terms] OR “Treatment Outcome”[MeSH Terms] OR

“Treatment Refusal”[MeSH Terms] OR “Stakeholder Participation”[MeSH Terms] OR “Patient Compliance”[MeSH Terms] OR “Health Resources”[MeSH Terms] OR

“Resource Allocation”[MeSH Terms] OR “Resource-Limited Settings”[MeSH Terms] OR “Delivery of Health Care”[MeSH Terms] OR

“health care quality, access, and evaluation”[MeSH Terms] OR “Healthcare Disparities”[MeSH Terms] OR “Health Workforce”[MeSH Terms] OR “Health Policy”[MeSH Terms] OR “Continuity of Patient Care”[MeSH Terms])

AND “Health Communication”[MeSH Terms]) OR “Health Facility Administration”[MeSH Terms] OR “Social Determinants of Health”[MeSH Terms]) AND (“Malawi”[MeSH Terms] OR “malawi*”[Title/Abstract])

### Embase search strategy: performed 24/08/2024

(’malawi’/mj OR ’malawi*’:ti,ab,kw)

AND

(’rheumatic heart disease’/mj OR ’rheumatic carditis’/exp OR ’rheumatic heart disease*’:ti,ab,kw OR ’rheumatic heart’:ti,ab,kw OR ’rheumatic carditis’:ti,ab,kw OR

(’rheumatic fever’/exp OR ’rheumatic fever*’:ti,ab,kw OR ’acute rheumatic fever*’:ti,ab,kw) OR

’streptococcal pharyngitis’/syn OR ’group a streptococcal infection’/de OR ’group a streptococcus’:ti,ab,kw OR ’group a streptococcal infection*’:ti,ab,kw)

AND

(’disease burden’/syn OR ’morbidity’/syn OR ’mortality’/syn OR ’prevalence’/de OR ’incidence’/de OR ’diagnosis’/syn OR ’patient referral’/exp OR ’therapy’/syn OR

’treatment outcome’/syn OR ’treatment refusal’/de OR ’patient compliance’/exp OR ’stakeholder engagement’/syn OR ’resource limited setting’/syn OR

’health care concepts’/exp OR ’health workforce’/syn OR ’health care policy’/de OR ’patient care’/syn OR ’hospital management’/syn OR ’social determinants of health’/syn)

### Cochrane Library Reviews search strategy: performed 24/08/2024

(rheumatic* OR “group A streptococcus” OR “rheumatic heart disease” OR “rheumatic fever” OR “acute rheumatic fever” OR “pharyngitis”)

AND

(“cost of illness” OR morbidity OR mortality OR prevalence OR incidence OR “healthcare access” OR “health care delivery” OR “health care quality” OR “resource allocation” OR “health care disparities” OR “health workforce” OR “patient compliance” OR “health policy” OR “social determinants of health”)

AND

Malawi

